# Antimicrobial resistance and mitigation strategies in healthcare settings: A scoping review

**DOI:** 10.1101/2020.07.02.20144915

**Authors:** O.O. Okeah, V. Morrison, J. Huws

**Affiliations:** Bangor University

## Abstract

**Background:** According to the European Center for Disease Prevention and Control (ECDC), the EU records an estimated 3.2 million healthcare associated infections (HAIs) and an associated 37,000 deaths annually. A significant proportion of the HAIs burden is attributable to multi-drug resistant organisms (MDROs). Infectious diseases remain top on the list of the leading causes of death globally with MDROs playing a significant role. Key amongst these organisms is *Clostridium difficile* and *Klebsiella pneumoniae* which belong to the broader group of ESKAPE pathogens.

**Aims:** This review aimed at identifying literature on interventions targeting *Clostridium difficile* and *Klebsiella pneumoniae*, their key outcomes, and the extent to which behavioural theory has been applied in such interventions.

**Methods:** This scoping review was undertaken and reported in accordance with the Preferred Reporting Items for Systematic Reviews and Meta-analysis Extension for Scoping Reviews (PRISMA-ScR) guidelines. The specific databases searched included MEDLINE, PubMed, Web of Science Core Collection, and CINAHL. The process for screening articles and data extraction was undertaken in duplicate by two reviewers. A narrative synthesis of the results is provided.

**Results:** The review included 34 studies (16 studies on *Clostridium difficile* and 18 articles focussed on *Klebsiella pneumoniae*). The specific antimicrobial stewardship interventions identified include **E**ducation, **S**urveillance and **S**creening, **C**onsultations, **A**udits, **P**olicies and **P**rotocols, **E**nvironmental measures, **B**undles of care, **I**solation precautions, as well as **N**otifications and alerts systems (ESCAPE-BIN). The identified outcomes include antimicrobial use, resistance rates, risk reduction, adherence to contact precautions, hospital stay, and time savings. Only one study incorporated Kotter’s stages of behaviour change and recorded the second largest (75%) sustained reduction in antimicrobials use whereas the remainder of the studies were devoid of behavioural approaches. The highest improvement (95%) in adherence isolation precautions was reported by an intervention involving the use of an IPC bundle and an environmental cleaning protocol.

**Conclusion:** This scoping review identified the available evidence on antimicrobial the mitigation strategies for *Clostridium difficile* and *Klebsiella pneumoniae* in healthcare settings as well as the key outcomes. There is need for further investigations on the feasibility of behaviour-based approaches in improving adherence of health workers to interventions targeting *Clostridium difficile* and *Klebsiella pneumoniae*.

## Introduction

Over the past centuries, infectious diseases have claimed millions of lives presenting a real threat to human existence^1^. The discovery of antimicrobial agents during the 19^th^ and 20^th^ century reduced the morbidity and mortality associated with infections^2^, and observations of Alexander Fleming on the effect of *Penicillium* on bacteria cultures birthed the era of anti-infective agents^3^. In 1947, Waksman, coined the term “antibiotic” in reference to a chemical agent capable of destroying or inhibiting the growth of microorganisms^4^. Progressively over the subsequent decades, clinicians recognised and employed antibiotics as an effective strategy for treating and eradicating pathogenic microorganisms.

As the use of antibiotics gained popularity worldwide with noted successes including the treatment of gram positive cocci with penicillin^3, 5^, a new threat namely antimicrobial resistance, emerged from the over-reliance on these life-saving therapeutic agents^6^. More than 50% of antimicrobials’ use is deemed as either inappropriate or unnecessary and within the last two decades alone, the use of antimicrobial agents has risen by 65% with available evidence confirming this as a key driver of antimicrobial resistance^7^. Coupled with the rapid human to human transmission of pathogens^8^, infectious microorganisms have continued to undergo adaptive evolution rendering a wide range of antimicrobial agents ineffective^9–11^. Consequently, infections such as tuberculosis have become even more potent as microorganisms continuously acquire resistance against previously effective antibiotics.

Today, infectious diseases remain top on the list of the leading causes of death globally based on recent statistics by the World Health Organization (WHO)^12^. Even more worrying are the deaths attributable to multi-drug resistant microorganisms (MDROs) that have continued to increase over the past decade. A modelling analysis reported 33,000 deaths associated with resistant bacteria in Europe in 2015, representing a significant rise since 2007^13^. Healthcare settings appear to have a higher risk for the human to human transmission of drug resistant pathogens. According to the European Center for Disease Prevention and Control (ECDC), the EU records an estimated 3.2 million healthcare associated infections (HAIs) and an associated 37,000 deaths annually^14^. The burden of HAIs within the EU translates to an estimated 2.5 million DALYs, 16 million additional hospitalization days, and an annual economic burden of 7 billion euros^15, 16^. A significant proportion of the HAIs burden is attributable to multi-drug resistant pathogens^17^.

Some of the multi-drug resistant pathogens associated with HAIs include ***E****nterococcus faecium, **S**taphylococcus aureus, **K**lebsiella pneumoniae, **A**cinetobacter baumannii, **P**seudomonas aeruginosa, and **E**nterobacter spp* acronymically referred to as ESKAPE pathogens^18–21^. In recent years, scientists have suggested the inclusion of *Clostridium (Clostridioides) difficile* also known as *C. diff* as a member of the ESKAPE pathogens and amending the acronym to **ESCAPE** pathogens^22^. For purposes of this study, we focused our attention on the research undertaken on *Clostridium difficile* and *Klebsiella pneumoniae* in healthcare settings.

*Clostridium difficile* accounts for the largest proportion of hospital-acquired diarrhoea attributable to the overuse of broad spectrum antibiotics that alter the profile of intestinal flora and trigger *Clostridium difficile* infections (CDIs)^23^. An European based study reported a 55% resistance rate of *Clostridium difficile* in isolates^23^. There is a large body of evidence confirming the transmission of *C. diff* within hospital environments^24, 25^, hence, the importance of proactive steps for mitigation.

On the other hand, a recent surveillance report on HAIs by the National Healthcare Safety Network reported a 9% prevalence of *Klebsiella pneumoniae* amongst hospitalized adult patients^17^ in European populations. This is a marginal increase considering a prior survey in the same population that reported an 8% prevalence of *Klebsiella pneumoniae* between 2011 and 2014^26^. A similar trend is evident in European paediatric intensive care units that recorded a 9% prevalence of *Klebsiella pneumoniae* between 2011 to 2014^27^.

### Surveillance and resistance patterns

#### Clostridium difficile

Various approaches are used in the surveillance of *Clostridium difficile* infections and resistance patterns. These methods include genomic analysis^28, 29^, polymerase chain reaction (PCR) ribotyping^30–32^, as well as molecular characterization of isolates^33–36^. Based on PCR-ribotyping, the resistant strains of *Clostridium difficile* examined by previous studies are mainly associated with ribotypes 012, 017, 018, 027, 053. 078, 176, and 630^37–43^.

Literature on the resistance patterns of *Clostridium difficile* reveals reduced susceptibility of the bacteria to fluoroquinolones^40, 44^, Macrolide-Lincosamide-Streptogramin B (MSLB) antibiotics^31^, erythromycin^45^, clindamycin^46^, moxifloxacin^47^, rifampicin^48^, rifamycin^49^, metronidazole^50, 51^, cadazolid^52^, linezolid^53^, imipenem^38^, vancomycin^54–56^, and fidaxomicin^54^. Studies have also shown that various factors contribute to *Clostridium difficile* resistance. These factors include wrong use of antibiotics^57^, intestinal microbiota^58^, overuse of antibiotics such as rifampicin in TB patients^48^, immunosuppression^59, 60^, gut dysbiosis^61^, and cancer treatment^61^.

#### Klebsiella pneumoniae

The risk factors for *Klebsiella pneumonia* resistance include hospitalization, recent antibiotics use, surgery, and renal failure^62, 63^ in adults whereas paediatric patients with a history of low birth weight, prolonged hospitalization, and prematurity are more susceptible to the resistant strains of *Klebsiella pneumoniae*^64^. The surveillance methods for *Klebsiella pneumoniae* include Whole Genome Sequencing (WGS), Metagenomic Sequencing^65–68^, isothermal DNA assays^69^, and pulsed field gel electrophoresis^70^.

There is demonstrable evidence of *Klebsiella pneumoniae* resistance against carbapenem, imipenem, meropenem, aminoglycosides, amoxicillin, amikacin, ampicillin/piperacillin, ciprofloxacin, levofloxacin, amoxicillin-clavulanic acid, trimethoprim-sulfamethoxazole, cefepime, colistin, nitrofurantoin, amikacin, aztreonam, ceftazidime, and tigecycline^71–80^.

Some of the resistant strains of *Klebsiella pneumoniae* include the clones ST11, ST29, ST101, ST258, which is also less susceptible to chlorhexidine cleaning, ST307, ST347, ST607-K25, ST661, ST1224, ST2558, ST3006^66, 81–86^. Whereas, the genes associated with resistance in *Klebsiella pneumoniae* species include *bla*_IMP 4,_ bla(OXA-48), OXA-33, TEM-1, and SHV-11, bla(KPC), bla(VIM), bla(NDM), wcaG, rmpA, intl1, blaCTX-M-15, qnrS1, qnrB1, aac(6’)-Ib, aac(6’)-Ib-cr, vagCD, *traT,* ccdAB, bla(CTX_M_1), bla(TEM), bia(OXA-1), *fyuA,* or *cnf-*1, and bla(SHV)^73, 81, 87–94, 95^.

### Rationale

Antimicrobial resistance (AMR) represents a public health emergency of global magnitude with the resultant mortality rate projected at 10 million fatalities by 2050^96^. The cost of treating resistant microorganisms has also significantly risen with evidence from the United States revealing a twofold increase between 2002 and 2015^97^. According to the World Health Organization, antimicrobial resistance is a preventable consequence of antibiotics’ misuse and overuse arising from a malfunctioned primary healthcare system^98^. The overuse of antibiotics is largely a prescription behaviour problem as healthcare professionals easily prescribe broad spectrum antibiotics without confirmatory laboratory tests causing the over flaring of *Clostridium difficile*. According to an England based study, the proportion of inappropriate antibiotics prescriptions in primary care trusts ranges between 8% and 23%^99^.

There is an urgent need to reduce the burden of AMR through multi-level approaches aimed at curbing transmission of multi-drug resistant organisms (MDROs), and optimizing the appropriate use of antibiotics. Although some efforts have been made to mitigate AMR^100^ including antimicrobial policies, the problem seems to persist given the high rates of inappropriate prescriptions^99,101^. Interventions for reducing transmission of MDROs have been encouraged and these are broadly categorized into horizontal measures and vertical measures^102^. The horizontal measures include pathogen non-specific strategies such as hand hygiene and environmental cleaning employed in disrupting the transmission^102^. The vertical measures are pathogen-specific and may include universal or targeted screening on admission for hospital care^103^. Additional strategies such as developing new antibiotics and exploring the possibility of effective vaccines could also potentially resolve the AMR issue^8^. Considering the potential implications, curbing the human-to-human transmission of pathogens and optimizing the use of antimicrobials appear to be practicable in most healthcare settings.

By focussing on *Clostridium difficile* and *Klebsiella pneumoniae,* we explored the widely researched topic of AMR with specific focus on the effectiveness of interventions targeting the drug resistant pathogens. Our preliminary exploration of literature retrieved three scoping reviews on antimicrobial misuse and interventions to address AMR. These scoping reviews had quite specific foci/targets and none addressed either *Clostridium difficile* or *Klebsiella pneumoniae.* The first scoping review^104^ was limited to dentistry settings; the second^105^ examined literature on knowledge, attitudes, and practices amongst community pharmacists and the third focussed on supply related interventions for reducing prescription of antibiotics in low-to-middle-income countries^100^. To expand the breadth of these reviews, we therefore scoped recent evidence on interventions for reducing *Clostridium difficile* and *Klebsiella pneumoniae* transmissions within wide ranging healthcare settings. Notably, we examined the volume of research on antimicrobial stewardship interventions aimed at optimizing antimicrobial agents against *Clostridium difficile* and *Klebsiella pneumoniae*.

### Research objectives

This scoping review addresses the question “What is the breadth of the available literature on interventions for reducing *Clostridium difficile* and *Klebsiella pneumoniae* transmission in healthcare settings?” and has the following objectives:

1. To identify existing literature on the interventions for reducing healthcare associated *C. difficile* and *Klebsiella pneumoniae* transmission.
2. describe the key outcomes for interventions targeting *Clostridium difficile* and *Klebsiella pneumoniae* in healthcare settings
3. assess the extent to which behavioural theory has been applied in interventions targeting *Clostridium difficile* and *Klebsiella pneumoniae* in healthcare settings

## Methods

Various methods exist for reviewing existing literature on research topics of interest namely systematic reviews, meta-analyses, and scoping reviews. Systematic reviews and meta-analyses are generally more applicable in contexts where the research question is narrow and focussed whereas scoping reviews are most preferable with broader research questions^106^ and where broadly mapping the literature for purposes of identifying main concepts, theoretical perspectives, available evidence, and gaps in literature is required^107^. Considering the broad nature of the present topic namely antimicrobial stewardship interventions in healthcare settings, the scoping review approach was justified.

Arksey and O’Malley proposed a five-stage framework for undertaking scoping reviews^108^. This staged approach is considered rigorous and enhances the transparency of the findings as sufficient detailing of the procedures employed at each stage allow for replication. The use of explicit approaches improves the reliability of the study and highlights the robustness of the employed methods^109^. The stages for conducting a scoping review entail the identification of a research question, identifying applicable studies, selecting studies for review, data charting, and collation of results, summarising, and compilation of reports^108^.

### Research protocol

Scoping review guidelines emphasize the importance of providing information regarding the protocol for purposes of improving transparency and minimising the risk for duplication^110^. The protocol for this scoping review is available on Open Science Framework (OSF) registries via https://osf.io/nk7wf.

This scoping review was undertaken and reported in accordance with the Preferred Reporting Items for Systematic Reviews and Meta-analysis Protocols (PRISMA-P) guidelines^110^. This approach integrates the five-stages proposed by Arksey and O’Malley with regard to the conduct of scoping reviews^108^. Subsequently, members of the research team reviewed this protocol prior to its online registration.

### Eligibility criteria

The review included peer-reviewed quantitative and/ experimental studies that either focus on reducing healthcare associated transmission of *C. diff* and *Klebsiella pneumoniae,* or on optimizing the use of antibiotics in relation to the aforementioned pathogens. Studies involving human participants published in English over the last ten years were included in this scoping review. Outbreak investigations that did not report any outcomes on the transmission of *C. diff* or *K. pneumoniae* as were studies that explored new diagnostic devices or therapeutic interventions with no outcomes on the hospital transmission of *C. diff* or *Klebsiella pneumoniae.* Table 1 below summarizes the eligibility criteria that was used to screen the retrieved articles.

**Table 1:** Eligibility criteria

|  | **Proposed criteria** | **Refined criteria** |
| --- | --- | --- |
| Population/<br>Setting | Healthcare facilities | Healthcare facilities |
| Intervention/<br>Exposure | AMS interventions for *C. diff* or *Klebsiella pneumoniae* | Optimizing use of antimicrobials or curbing transmission of *C. diff* and/ or *Klebsiella pneumoniae* |
| Comparison | No intervention | No intervention |
| Outcome | Control of *C. diff* and/ or *Klebsiella pneumoniae* | Appropriate use of antimicrobials<br>Reduced risk/ transmission of *C. diff* and/ or *Klebsiella pneumoniae* |
| Study designs | All study designs | Observational studies, quasi-experimental studies, randomised controlled trials (RCTs) |

### Information sources

It is recommended that the search for existing literature should be undertaken comprehensively across various platforms in order to realise the purpose of a scoping review^108, 110, 111^. For purposes of this review, the search for literature spanned across electronic databases accessible through the Bangor University library search engine, bibliographies, key journals, and websites for relevant organisations.

The specific databases searched included MEDLINE via EBSCO*host*, PubMed Open Access via NCBI, Web of Science Core Collection, and CINAHL Plus via EBSCO*host*. The institutional databases explored for literature search included the World Health Organisation, the British Society for Antimicrobial Chemotherapy, as well as the National Institute for Health Research.

### Literature search

The search for sources was undertaken with the assistance of the Bangor University librarian. Appendix 1 provides details of the search strategy applied across the databases.

### Study selection

Two reviewers (BO and JH) independently applied the inclusion and exclusion criteria on the retrieved articles for inclusion in this review. Full articles for studies that provided a best fit to the central research question were retrieved and the reviewers read the full texts to determine their inclusion in the review. A mechanism for discussing disagreements with a third reviewer (VM) and consensus building was in place.

### Data charting

The data extraction form should be as comprehensive as possible for the charting of relevant data from the identified evidence sources^108, 111^. Existing guidelines also recommend detailing the process of developing, calibrating the charting form, the charting process, as well as the resolution of disagreements^110^. It is also important to update the charting form iteratively with descriptions of any revisions for improved transparency.

The research team developed a form for abstracting data in order to capture all the relevant variables from the identified sources. The standardized form allowed for extraction of the main study characteristics as well as the specific metrics relevant to the central research question of this scoping review. The form was subjected to preliminary calibration to ensure its accuracy, consistency, and reliability.

The data items extracted from each study included the reference, the study type, the study objectives, population or setting, country, the intervention, intervention duration, healthcare workers involved, outcome measures or findings, and the conclusions of the study.

### Results collation, summary, and report compilation

The guidelines for scoping reviews recommend that reviewers provide a comprehensive overview of the retrieved evidence^108^. This involves organising the evidence based on the identified themes and giving a narrative account as opposed to synthesising the results. Scoping reviews are useful in mapping relevant concepts that underpin a phenomenon of interest while aggregating the existing evidence on the topic^110, 111^. The subsequent sections provide a narrative synthesis of the existing literature on AMS interventions for *C. diff* and *Klebsiella pneumoniae* as well as any apparent gaps in line with the central review question.

## Results

### Selection of sources of evidence

**Figure 1:**
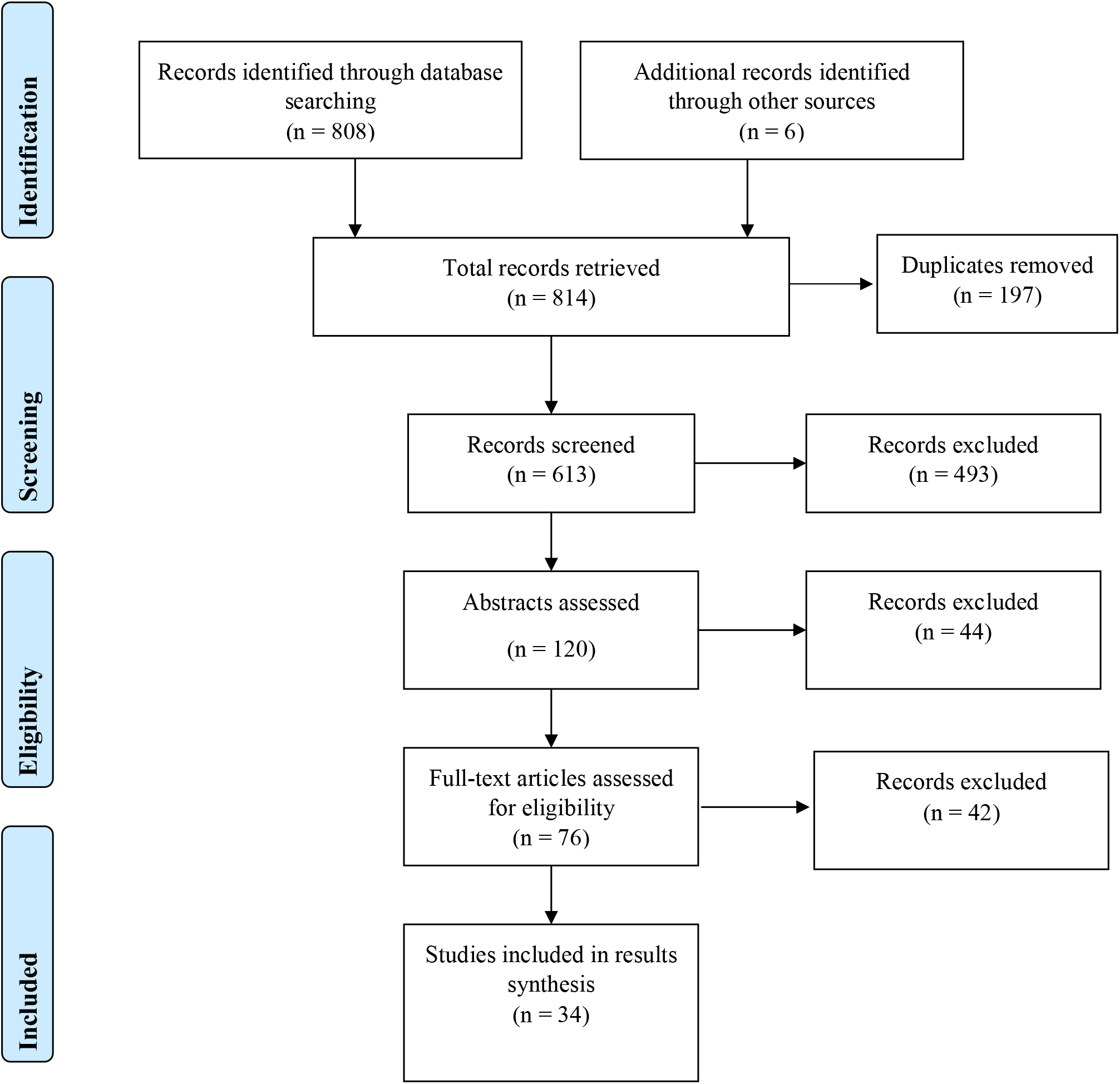
PRISMA flow diagram

The search for literature across the identified databases (MEDLINE, CINAHL, PubMed, and Web of Science) retrieved 808 records whereas an additional six titles were identified through bibliographic searches. Following de-duplication, 613 titles were screened where 493 records were considered no to meet the inclusion criteria. The abstracts of the remaining 120 records were further screened leading to the exclusion of 44 articles. Full text reading was done for 76 articles leading to the exclusion of 42 articles and inclusion of 34 articles.

### Characteristics of selected studies

16 studies (see Table 2) focussed on *Clostridium difficile*^112, 113, 114–121, 122–127^ and 18 studies (see Table 3) focussed on *Klebsiella pneumoniae*^128, 129, 130–137, 138–145^. The studies varied in their designs with majority being quasi-experiments (31 articles). The other study designs included cohort studies (2 articles) and a secondary analysis of a randomized controlled trial (1 article). 27 of the studies were undertaken prospectively whereas 7 studies followed a retrospective approach.

**Table 2:** Clostridium difficile interventions

|  | References |  |  |  |  |  |  |  |  |  |  |  |  |  |  |  |
| --- | --- | --- | --- | --- | --- | --- | --- | --- | --- | --- | --- | --- | --- | --- | --- | --- |
| Interventions | 112 | 113 | 114 | 115 | 116 | 117 | 118 | 119 | 120 | 121 | 122 | 123 | 124 | 125 | 126 | 127 |
| Surveillance/ Screening |  |  |  |  | ✓ |  | ✓ |  |  |  |  | ✓ |  |  |  |  |
| Alerts and notifications |  |  |  |  |  |  | ✓ |  |  |  |  | ✓ |  |  | ✓ |  |
| Isolation precautions |  |  |  |  |  |  | ✓ |  |  |  |  | ✓ |  | ✓ | ✓ | ✓ |
| Environmental disinfection |  |  |  |  |  |  |  |  |  | ✓ |  |  |  | ✓ |  | ✓ |
| Audits and feedback | ✓ |  | ✓ | ✓ |  | ✓ |  |  |  |  | ✓ | ✓ | ✓ |  |  |  |
| Consultations |  | ✓ | ✓ |  |  |  |  |  |  |  |  |  |  |  |  |  |
| Antimicrobial policies and/<br>protocols |  | ✓ |  |  | ✓ |  |  |  |  |  | ✓ |  |  |  | ✓ |  |
| Care bundles |  |  |  |  |  |  |  |  |  |  |  |  |  |  |  | ✓ |
| Staff education |  | ✓ | ✓ |  | ✓ |  |  |  |  |  | ✓ |  |  | ✓ |  |  |
| Biocidal (Cu<sub>2</sub>O) linen |  |  |  |  |  |  |  | ✓ | ✓ |  |  |  |  |  |  |  |
| Intervention duration (months) | 24 | 18 | 18 | 12 | 16 | 18 |  | 8 | 27 | 27 | 16 | 12 | 13 | 24 |  | 22 |

**Table 3:** Klebsiella pneumoniae interventions

|  | References |  |  |  |  |  |  |  |  |  |  |  |  |  |  |  |  |  |
| --- | --- | --- | --- | --- | --- | --- | --- | --- | --- | --- | --- | --- | --- | --- | --- | --- | --- | --- |
| Interventions | 128 | 129 | 130 | 131 | 132 | 133 | 134 | 135 | 136 | 137 | 138 | 139 | 140 | 141 | 142 | 143 | 144 | 145 |
| Surveillance/ Screening | ✓ | ✓ |  |  |  | ✓ | ✓ | ✓ | ✓ | ✓ | ✓ | ✓ | ✓ | ✓ | ✓ |  | ✓ | ✓ |
| Alerts and notifications | ✓ |  | ✓ | ✓ |  | ✓ |  | ✓ | ✓ |  |  |  |  |  |  |  |  |  |
| Isolation precautions | ✓ |  | ✓ |  |  | ✓ | ✓ | ✓ | ✓ | ✓ | ✓ | ✓ | ✓ | ✓ | ✓ | ✓ | ✓ |  |
| Environmental disinfection |  |  | ✓ |  |  |  | ✓ |  | ✓ | ✓ |  | ✓ | ✓ | ✓ |  |  |  |  |
| Audits and feedback |  |  |  | ✓ |  | ✓ |  |  |  | ✓ |  |  |  | ✓ |  | ✓ |  |  |
| Consultations |  |  |  |  |  |  | ✓ |  |  |  |  |  |  |  |  |  |  |  |
| Antimicrobial policies and protocols | ✓ |  |  | ✓ | ✓ |  |  |  |  |  |  |  |  |  |  |  | ✓ |  |
| Care bundles |  |  |  |  |  |  |  |  |  |  |  | ✓ |  |  |  |  |  |  |
| Staff education and/ patient education |  |  | ✓ | ✓ |  |  |  |  |  | ✓ |  |  |  | ✓ |  | ✓ |  |  |
| Intervention duration (months) | 36 | 14 | 48 | 36 | 24 | 14 | 2 | 36 | 6 | 17 | 4 | 24 | 8 | <1 | 2 | 72 | 3 | 6 |

32.4% (11) of the studies were conducted in the United States of America 115,118,141,146,119–122,124,126,127,136 whereas two studies each are based in Canada^112, 114^ and Greece^135, 137^. Four of the retrieved studies are from Italy^113, 117, 134, 142^ while Israel^125, 130, 133^ and China^129, 139, 140^ had three studies each. Lastly, the selected articles included one study each from Japan^123^, United Kingdom^116^, South Africa, Denmark^131^, Brazil^132^, France^138^, South Korea^143^, Hungary^144^, and the Netherlands^145^.

Most of the studies (30 articles) were single site studies whereas four studies are multi-site experiments^112, 119, 121, 127^. There were variations in the study populations with three studies on *Klebsiella pneumoniae* involving neonates in the neonatal intensive care unit^140, 141, 144^ whereas 31 studies involved adult subjects admitted for care within the hospital settings. All the studies on *Clostridium difficile* were based on adult populations probably due to the evidence supporting advanced age as a risk factor for CDIs while three interventions targeting *Klebsiella pneumonia* involved neonatal populations^140, 141, 144^.

### Synthesis of results

#### Interventions

The interventions varied across the included studies and either targeted the use of antimicrobial agents or interrupting the transmission of *Clostridium difficile* and *Klebsiella pneumoniae* using additional IPC measures within the hospital environments. The duration of interventions varied across the studies from three weeks^141^ up to six years^143^. The interventions involved various cadres of professionals namely infectious disease (ID) experts^113, 114, 124, 125, 139^, consultants^117, 130, 139, 141, 145^, nurses^113, 125, 127, 128, 130, 133, 134, 137–139, 140, 141, 144, 145^, doctors^116, 123–125, 137^, physicians^112, 113, 115, 117, 118, 122, 124, 127, 128, 133, 138, 139, 141, 143, 145^, pharmacists^112, 115–117, 122, 124–126^, epidemiologists^125, 128, 136, 141^, laboratory personnel^133^, microbiologists^124, 133, 138, 141^, and support staff (cleaners, caregivers, housekeepers, paramedics, porters, environmental officers)^125, 127, 130, 134, 141, 144, 145^. Additional cadres involved include managers^116, 141^, infection control staff^118, 123, 127, 128, 130, 133, 134, 140, 141^, unspecified clinicians/ medical personnel^118–121, 123, 134–137, 138, 142–144^, quality improvement (QI) staff^124^, patients^130^, public health (PH) staff^133^, and patient visitors^145^. The bar graph below summarizes the proportions of studies that involved various health professionals.

**Figure 2:**
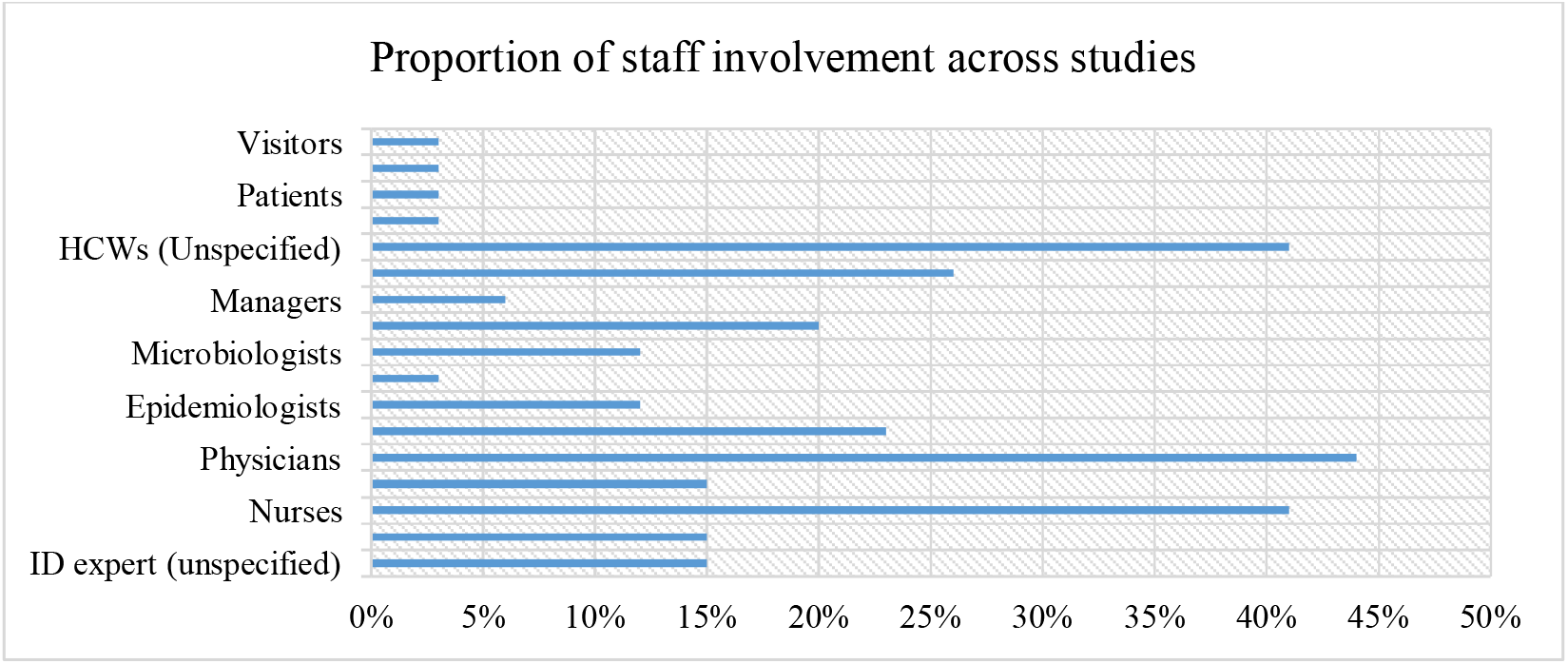
Proportion of staff involvement across studies

Most of the interventions were multi-faceted involving the implementation of at least two strategies to achieve the intended outcomes as highlighted in Table 2. The strategies employed in interventions targeting *Clostridium difficile* included surveillance and/ or active screening^116, 118, 123^, alerts and notifications^118, 123, 126^, isolation precautions^118, 123, 125–127^, environmental disinfection^121, 125, 127^, audits and feedback^112, 114, 115, 117, 122–124^, antimicrobial policies and/ protocols^113, 116, 122, 126^, care bundles^127^, staff education^113, 114, 116, 122, 125^, and specialised biocidal linen^119, 120^. Table 2 below outlines the combination of these interventions across the identified studies.

The commonest strategy targeting *Clostridium difficile* reported across seven studies involved the use of audits and feedback^112, 114, 115, 117, 122–124^. This entailed reviewing the prescribed antibiotics by an antimicrobial pharmacist^112, 114, 115, 117, 122, 124^ or the infection control team^123^ and providing feedback to the prescriber for further action. In some instances, the audits were undertaken offsite using electronic records systems^114, 115^ and teleconferences. Audits were also combined with staff education sessions organised on identified gaps aimed at optimising the use of antimicrobials^114, 122^. Some interventions combined the audits with restrictive antimicrobial policies and review of treatment protocols occasionally requiring approval prior to the use of a targeted antibiotic^122^. Another intervention combined audits with screening patients and notifying physicians on detection of *C. difficile*, promptly isolating infected patients, and monitoring appropriate use of antibiotics with prompt feedback to the responsible doctors^123^.

Additional interventions with a component of staff education included bedside infectious diseases consultation^113^, restrictive antimicrobial policies and protocols^113, 116, 125^, and contact precautions^125^. Bedside consultations involved a part-time infectious diseases expert reviewing patients on antibiotic treatment three times a week and discussing the same with attending physicians^113^. This was coupled with revising antimicrobial treatment protocols and educating staff on reducing the inappropriate use of antimicrobials^113^. The second intervention involved educating prescribers coupled with restriction and eventual abolishment of two broad-spectrum antibiotics namely ceftriaxone and ciprofloxacin from wards^116^. Lastly, an intervention undertaken in a geriatric hospital involved educating all healthcare workers on isolation precautions and environmental disinfection as well as policies restricting the use of antibiotics^125^.

Isolation precautions were also incorporated in other interventions including a multi-site collaborative intervention involving an infection prevention bundle to encourage adherence to contact precautions and an environmental cleaning protocol^127^. The isolation precautions included nursing patients in a single room, hand washing at recommended times, and the use of appropriate personal protective equipment namely gloves, and disposable aprons. Environmental decontamination entailed the use of appropriate decontamination agents to clean the patient environment and reduce the presence of *Clostridium difficile.* A single centre study combined isolation precautions with a computer generated real time notification system for toxigenic *C. difficile* results and a treatment protocol using vancomycin only or vancomycin and metronidazole^126^. The final study on isolation precautions also incorporated an automated system that tracked *C. difficile* results and triggered alerts on the patient’s electronic records as well as automatically ordering for the appropriate isolation precautions thus aiding the healthcare personnel’s actions^118^.

Three standalone interventions targeted modifying the hospital environment and reduce the bioavailability of *Clostridium difficile*^119–121^. A multisite randomised controlled trial employed four disinfection strategies for environmental cleaning following the discharge of *C. difficile* and other MDROs patients^121^. These strategies included standard disinfection with an ammonium solution or 10% hypochlorite (bleach), standard disinfection with ultraviolet (UV) light or bleach with UV light, bleach only, or UV light with bleach^121^. Lastly, two quasi-experiments involved replacing hospital linen with coper oxide impregnated bedsheets, pillow cases, washcloths, and towels^119, 120^. This is because of copper’s biocidal activity on some drug resistant bacteria including *Clostridium difficile*.

Interventions targeting the containment or reduced transmission of *Klebsiella pneumoniae* included surveillance and/ or active screening^128, 129, 133–140, 141, 142, 144, 145^, alerts and notifications^128, 130, 131, 133, 135, 136^, isolation precautions^128, 130, 133–140, 141, 142, 144^, environmental decontamination^130, 134, 136, 137, 139–141^, antimicrobial audits and feedback^131, 133, 137, 141^, specialist consultations^134^, antimicrobial policies and/ or protocols^128, 131, 132, 144^, care bundles^139^, and staff and/ or patient education^130, 131, 137, 141, 142^.

The commonest strategy targeting *Klebsiella pneumoniae* appears to be surveillance or active screening and cultures to detect the presence of *K. pneumoniae*. One surveillance intervention involved the use of a flagging system for suspected patients at the emergency department, cohorting cases, sampling cultures from healthcare hands of personnel and the environment, and a policy restricting use of carbapenems^128^. Another multisite intervention combined routine screening of patients with mandatory isolation of confirmed cases including having dedicated staff looking after patients coupled with a requirement to notify all carbapenem resistant cases to public health authorities^133^. Similarly, a surveillance intervention in a 250-bed general hospital required adherence to isolation precautions and compulsory notification of public health authorities on identified cases^135^.

An outbreak containment intervention in an ICU setting employed active screening of patients, disinfection of the environment and respiratory equipment, and isolation precautions. One standalone intervention investigated the effectiveness of active screening of patients on the detection of *Klebsiella pneumoniae* in an ICU setting^129^ while another study tracked sporadic hospital outbreaks using whole genome sequencing^145^. An observational study relied on rectal swabs for the active surveillance of *Klebsiella pneumoniae* in a cancer centre and a tertiary hospital^136^. Subsequently, the confirmed cases were promptly isolated requiring healthcare personnel’s adherence to contact precautions and environmental cleaning protocols^136^. Other surveillance intervention similarly effected isolation precautions for confirmed cases^138^ combined with either environmental cleaning protocols, staff education, adherence audits, or a bathing protocol^137, 138, 140–142, 144^. An intervention based in an Israeli medical centre rolled out isolation guidelines in combination with staff education, environmental cleaning protocols supported with a computerized system for flagging CRKP cases^130^. A multi-disciplinary intervention in a 510-bed Danish university hospital employed Kotter’s eight stages of change by delivering staff training and use of notification systems to enhance isolation precautions, and appropriate use of antimicrobial agents^131^. An antimicrobial stewardship intervention in a Brazilian tertiary care hospital examined the effectiveness of a restrictive antimicrobial policy on the use of carbapenems^132^. Lastly, a south-Korean based study in a 900-bed tertiary university hospital examined the examined the effectiveness of enhanced contact isolation precautions on CRKP incidence. This was delivered through staff education, auditing prescriptions and discontinuing inappropriate antibiotics within 72 hours, and strict adherence to contact precautions including hand hygiene, single use gowns, and gloves.

#### Outcomes

The key outcome measures reported across the studies included consumption of targeted antimicrobials and/ or associated costs^112, 113, 114–117, 122, 123, 128, 131, 132^, incidence of *Clostridium difficile*^112, 113, 114–117, 119–122, 123–125, 127^ or incidence and/ resistance rates of *Klebsiella pneumoniae*^128, 129, 130–134, 137, 139, 143, 144^ as well as risk on other HCAIs^114, 121, 123, 127, 130, 137, 139^. Additional outcomes included containment of an outbreak of the targeted organisms^135, 136, 138, 140–142, 145^, adherence to isolation precautions^114, 120, 127, 134– 136, 140, 143, 144^, time savings^118, 126^, hospital stay^144^, and mortality^113^.

The use of antibiotics was measured in daily defined doses (DDD) per patient population before and after the intervention^112–114, 116, 117, 122, 123^. Occasionally, some interventions reported on the proportionate changes on the costs of antibiotics^114, 117, 124^ as well as resistance rates. The risk measures for either *C. difficile* or *K. pneumoniae* as well as other HCAIs reported in number of cases per population before and after the intervention and converted to absolute risk (%). Another outcome measure reported by some studies was the proportionate reduction in treatment times, hours saved by healthcare workers per number of admissions, or proportionate reduction in the duration of hospital stay. Adherence to isolation precautions was measured based on the proportion of staff members that complied with hand hygiene guidelines, use of disposable aprons and gloves, and environmental cleaning. Lastly, mortality outcomes related to the number of deaths attributed to either *Clostridium difficile* or *Klebsiella pneumoniae*. The outcome measures for *Clostridium difficile* and *Klebsiella pneumoniae* interventions are summarised in Table 4 and Table 5 respectively.

**Table 4:** Summary of outcomes for Clostridium difficile interventions

| Key outcomes | References |  |  |  |  |  |  |  |  |  |  |  |  |  |  |  |
| --- | --- | --- | --- | --- | --- | --- | --- | --- | --- | --- | --- | --- | --- | --- | --- | --- |
|  | 112 | 113 | 114 | 115 | 116 | 117 | 118 | 119 | 120 | 121 | 122 | 123 | 124 | 125 | 126 | 127 |
| Antimicrobials use (DDD/1000PDs) | ↓310 | ↓200 |  | ↓6.58 | ↓124 | ↓141 |  |  |  |  | ↓34 | ↓10.7 |  |  |  |  |
| Antimicrobials use (% reduction) | 11 | 47 | 46 |  | 72.5-95 | 22 |  |  |  |  | 12 | 37 |  |  |  |  |
| Antibiotics cost (↓%) |  |  | ↓54 |  |  | ↓24 |  |  |  |  |  |  | ↓51 |  |  |  |
| Antimicrobials streamlining (%/week) |  |  |  |  |  |  |  |  |  |  |  |  | ↑52 |  |  |  |
| Resistance rates |  |  |  |  |  |  |  |  |  |  |  |  |  |  |  |  |
| CD risk/100,000 or/10000PDs (post-intervention) | ⇔ | 12 |  | 14 | 55 | 60 |  |  | 2.8 | 170 | 2.8 | 11 | 16 | ⇔ |  | 85 |
| CD absolute risk (%) | ⇔ | ↓67 | ↓46 | ↓83 | ↓77 | ↓31 |  | ↓51 | ↑87 | ↓5 | ↓71 | ↓36 | ↓71 | ⇔ |  | ↓37 |
| Risk for HCAs (AR) |  | ↓25 |  |  | 17-25 |  |  |  |  | ⇔ |  | ↓ |  |  |  | ↓ |
| % reduction in time for start of treatment |  |  |  |  |  |  |  |  |  |  |  |  |  |  | 64 |  |
| Time savings (hrs/1000 admissions) |  |  |  |  |  |  | ↓43 |  |  |  |  |  |  |  |  |  |
| Hospital stay |  |  |  |  |  |  |  |  |  |  |  |  |  |  |  |  |
| Adherence to isolation precautions (%) |  |  |  |  |  |  |  |  | ↓6 |  |  |  |  |  |  | ↑95 |
| Mortality |  | ⇔ |  |  |  |  |  |  |  |  |  |  |  |  |  |  |

DDD: Daily defined doses; PD: Patient days; CD: *Clostridium difficile*; HCAs: Healthcare associated infections; AR: Absolute risk, ↓: Reduced ↑: Increases; ⇔: Remained the same; ●: Outbreak was contained.

**Table 5:** Summary of outcomes for Klebsiella pneumoniae interventions

| Key outcomes | References |  |  |  |  |  |  |  |  |  |  |  |  |  |  |  |  |  |
| --- | --- | --- | --- | --- | --- | --- | --- | --- | --- | --- | --- | --- | --- | --- | --- | --- | --- | --- |
|  | 128 | 129 | 130 | 131 | 132 | 133 | 134 | 135 | 136 | 137 | 138 | 139 | 140 | 141 | 142 | 143 | 144 | 145 |
| Antimicrobial use (DDD/1000PDs) | ↓ |  |  |  | 13 |  |  |  |  |  |  |  |  |  |  |  |  |  |
| Antimicrobials use (% reduction) |  |  |  | ↓75 | ↓21 |  |  |  |  |  |  |  |  |  |  |  |  |  |
| Antibiotics cost (↓%) |  |  |  |  |  |  |  |  |  |  |  |  |  |  |  |  |  |  |
| Antimicrobials streamlining (%/week) |  |  |  |  |  |  |  |  |  |  |  |  |  |  |  |  |  |  |
| Resistance rates |  | ↓ |  |  | ↔ |  |  |  | ● |  |  |  |  |  |  | ↓ |  |  |
| KP risk/100,000 or/10000PDs | 18 |  | 0.5 | 23% |  | ↓ | 56 |  | ↓ | ↓ | ● | 28 |  |  |  | 0.9 | ↓ |  |
| KP absolute risk (%) | ↓97 |  | ↓92 | ↓17 |  | 12 | 12 | ● | ● | 10 | ● |  | ● | ● | ● | 46 | ↓ | ● |
| Risk for HCAs (AR) |  |  | ↓55 |  |  |  |  |  |  | ↑59 |  | ↓84 |  |  |  |  |  |  |
| % reduction in time for start of Rx |  |  |  |  |  |  |  |  |  |  |  |  |  |  |  |  |  |  |
| Time savings (hrs/1000 admissions) |  |  |  |  |  |  |  |  |  |  |  |  |  |  |  |  |  |  |
| Hospital stay (%PDs) |  |  |  |  |  |  |  |  |  |  |  |  |  |  |  |  | ↓15 |  |
| Adherence to isolation precautions (%) |  |  |  |  |  |  | ↑ | ↑ | ↑ |  |  |  | ↑ |  |  | ↑35 | ↑29 |  |
| Mortality |  |  |  |  |  |  |  |  |  |  |  |  |  |  |  |  |  |  |

DDD: Daily defined doses; PDs: Patient days; HCAs: Healthcare associated infections; AR: Absolute risk, ↓: Reduced ↑: Increases; ↔: Remained the same; ●: Outbreak was contained

#### Clostridium difficile

##### Antimicrobial use

Seven studies reported variations in the consumption of antimicrobial agents following the stewardship interventions^112, 114, 115, 117, 122–124^. The changes in antimicrobial use are reported in daily defined doses per 1000 patient days (DDD/1000PDs). Reduced use of antimicrobials ranged between 6.58 DDDs/1000 PDs and 310 DDDs/1000 PDs.

The largest absolute reduction in antimicrobial use of 310 DDs/1000PDs was reported from an intervention that involved audits and feedback systems^112^. However, this reduction was the least in proportionate terms (11%) compared to the largest proportional reduction in antimicrobials use of 79% after an intervention involving restrictive antimicrobial policies and staff education^116^.

In terms of antimicrobial costs, the largest reduction in expenditure (54%) was reported from intervention involving half-hour monthly staff education sessions and audits of prescribed antibiotics using a structured electronic checklist^114^. 679 patients from two internal medicine units in a tertiary care hospital were observed over an 18 months period^114^. One study reported a 52% improvement in antimicrobial streamlining after undertaking weekly reviews of prescribed antibiotics combined with remote consultations with an infectious diseases pharmacist through teleconferencing^124^. The latter study was conducted in a 141-bed community hospital over 13 months^124^. None of the *C. difficile* targeting interventions reported on the resistance rates for specific antimicrobial agents following their implementation.

##### Risk for CDIs, other HCAIs, and associated mortality

Fourteen studies reported the impact of the interventions on the risk for CDIs or other healthcare associated infections^112, 113, 114–117, 119–122, 123–125, 127^. The highest overall reduction of 83% in absolute risk for CDIs was reported from a 12-months audits and feedback intervention involving physicians and pharmacists and pharmacists in a 212-bed Massachusetts hospital^115^. On the other hand, a 24-months multisite intervention amongst leukemia patients involving audits and feedbacks^112^ reported no impact on the risk of CDIs and associated mortality. Similarly, a second 24-months cross-sectional study involving geriatric patients from two Israeli hospitals that entailed staff education, environmental disinfection, and isolation precautions had no impact on CDIs^125^.

Regarding the effect of CDI interventions on other HCAIs, an antimicrobial stewardship intervention in a 150-bed spinal injury hospital involving bedside infectious diseases consultation, staff education, and antimicrobial policies reported a 25% absolute risk reduction for other HCAIs^113^. This is also the only study that reported on CDI associated mortality whereby no differences were observed between the experimental and control groups^113^. A multisite RCT investigating the effectiveness of four environmental disinfection strategies reported no effect on the risk of other HCAIs^121^. A third study assessing the impact of intensified IPC precautions on MDROs implemented over twelve months in a 409-bed Japanese tertiary hospital simply indicated there was a reduction in the risk of other HCAIs^123^.

Two studies involving the use of biocidal linen impregnated with copper oxide reported contradictory findings which could be partly due to the differences in study settings. The first study involved six hospitals in both urban and rural settings with 1019 beds in total implemented over eight months (568,397 patient days) and reported a 51% reduction in the risk for CDIs^119^. The second study was undertaken in a single long-term care hospital over 27 months (29,342 patient days) reported an 87% increase in the risk of CDIs^120^. In the latter study, the researchers also acknowledged that study participants were never blinded possibly leading to the deterioration of contact precautions specifically hand hygiene that reduced by 6%^120^.

##### Adherence to isolation precautions

The highest improvement (95%) in adherence to isolation precautions was reported by a 22-months multisite (35 hospitals) intervention involving the use of an infection prevention bundle with isolation precautions and an environmental cleaning protocol^127^. On the other hand, an intervention involving the use of biocidal linen impregnated with copper oxide reported a 6% reduction in adherence to isolation precautions^120^ as previously discussed.

##### Time savings

Two studies reported outcomes related to time savings^118, 126^. The first intervention involved treatment protocols for *C. difficile*, real-time computerized notifications of toxigenix *C. difficile results*, and isolation precautions. This was undertaken in a 433-bed adults medical center and recorded a 64% reduction in time prior to the initiation of appropriate antibiotics treatment^126^. The second study involving active surveillance, an alert system, and isolation precautions in a 410-bed hospital treating trauma, burns, and cancer patients reported a 43% reduction in care hours per 1000 admissions^118^. There were no studies on *C. difficile* that assessed whether the interventions affected the length of hospital stay.

#### Klebsiella pneumoniae

##### Antimicrobials use

Three studies reported on antimicrobial use with regards to *Klebsiella pneumoniae* interventions^128, 139, 140^. One study involving a flagging system for confirmed cases, isolation precautions, and a carbapenems restriction policy in a 1000-bed tertiary university hospital simply indicated there was a reduction in the use of meropenem^128^. The second study employed Kotter’s stages of change in a multi-disciplinary intervention involving staff education, notifications on prescription of restricted antibiotics and antimicrobial protocols in a 510-bed Danish hospital recorded a 75% reduction in the use of targeted antibiotics^131^. The last study involving restrictive antimicrobial policies reported a 21% (12.9 DDDs/1000 PDs) reduction in the use of targeted antimicrobial agents^132^. Four (22%) studies reported on the resistance rates for specific antibiotics associated with *K. pneumoniae resistance* either as a reduction^129, 143^ or no effect^132, 136^ on the resistance rates with no absolute figures on the same. No intervention was associated with a reciprocal increase in the antibiotics resistance rates of *Klebsiella pneumoniae*.

##### Risk for Klebsiella pneumoniae, other HCAIs, and associated mortality

Seven studies reported on containment of Klebsiella pneumoniae outbreaks with no outcomes on the residual risk for the bacteria^135, 136, 138, 140–142, 145^. Two interventions involving active surveillance through screening^129^ and staff education combined with isolation precautions^143^ reported a reduction in the resistant rates of Klebsiella pneumoniae. The first intervention was conducted over 14 months in an ICU setting in China^129^ while the second intervention was undertaken in a 900-bed tertiary hospital in South Korea^143^. A 24-months intervention in a tertiary hospital (200 beds) involving restriction of group 2 carbapenems for gram negative bacteria recorded no changes in the resistance rates of Klebsiella^132^.

Regarding the absolute risk of Klebsiella pneumoniae, the largest risk reduction (97%) the 36-months hospital wide intervention described above aimed at eradicating carbapenem resistant *Klebsiella pneumoniae* (CRKP)^128^. This intervention involved physicians, epidemiologists, nurses, and the infection control team. The lowest reported reduction in the absolute risk of *Klebsiella pneumoniae* was from a 17-months multi-faceted intervention that entailed active surveillance, isolation precautions, audits and feedback, environmental cleaning, and staff education^137^.

Another intervention involving staff education, isolation, environmental cleaning, computerized flagging of cases reported a 55% reduction in other HCAIs^130^ while a second intervention comprising of screening, isolation, environmental disinfection, and care bundles reported an 84% reduction in other HCAIs over a 48 months period^139^. On the other hand, one study reported a 59% rise in the risk of other HCAIs following an intervention that involved screening, isolation, environmental decontamination, audits, and education over a 17 months duration^137^. The intervention involved 601 patients retrospectively and 250 patients prospectively in the solid organ transplant (SOT) department. The increase in the incidence of other carbapenem resistant organisms was attributed to the intrahospital transfer of carriers to the SOT department and the subsequent transfer of post-surgical patients to the ICU where they were allegedly colonized by bacteria^137^. There are no studies that reported on the impact of interventions on mortality associated with *Klebsiella pneumoniae* in healthcare settings.

##### Hospital stay and adherence to contact precautions

Only one study recorded outcomes associated with hospital stay whereby there was a 15% reduction in the hospitalization duration that was also associated with 29% increase in adherence with contact precautions^144^. This three-months intervention involved 355 patients in a 17-bed neonatal intensive care unit in Hungary^144^. Another six years intervention involving staff education also reported a 35% improvement in adherence to contact precautions^143^. Lastly, four additional studies also reported an improvement in adherence to contact precautions although this was not reported in numerical values^134–136, 140^.

## Discussion

### Summary of evidence

In this scoping review, we identified studies on antimicrobial stewardship interventions for *Clostridium difficile* and *Klebsiella pneumoniae* in healthcare settings published between 2010 and 2019. The first set of interventions focussed on optimal use of antimicrobial agents and included restrictive antimicrobial policies and treatment protocols, specialists’ consultations, notifications and alert systems, as well as audits and feedback (also referred to as academic detailing). The second set of interventions aimed at curbing the healthcare associated transmission of *Clostridium difficile* and *Klebsiella pneumoniae* included surveillance and active screening, isolation precautions, environmental disinfection, use of care bundles, and education of staff and or patients. There was an additional intervention specific to *Clostridium difficile* namely the use of biocidal linen impregnated with copper oxide which can be grouped as part of the environmental modification measures.

We propose the abbreviation ESCAPE-BIN (**E**ducation, **S**urveillance/**S**creening, **C**onsultations, **A**udits, **P**olicies and **P**rotocols, **E**nvironmental measures, **B**undles of care, **I**solation, and **N**otifications or alerts) to denote these cross-cutting interventions for disrupting the transmission cycle of *Clostridium difficile* and *Klebsiella pneumoniae* in healthcare settings. As discussed previously discussed in this paper, *Clostridium difficile* and *Klebsiella pneumoniae* belong to the wider group of ESKAPE pathogens with common modes of patient-to-patient transmission in healthcare settings such as contact transmission by healthcare workers. The findings above also show that interventions targeting either *Clostridium difficile* and *Klebsiella pneumoniae* have a significant impact on the health care associated risk of other ESKAPE pathogens with similar modes of transmission. As such, strategies like improving adherence to contact precautions targeting any of the organisms could potentially reduce the spread of other pathogens within the same setting. The acronym **ESCAPE-BIN** may therefore be applicable in denoting these interventions for limiting the healthcare transmission of ESKAPE pathogens.

Our review found that most interventions targeting *Clostridium difficile* tend to integrate a component of restrictive antimicrobial policies or treatment protocols. On the other hand, interventions targeting *Klebsiella pneumoniae* mainly incorporated screening, isolation precautions, or environmental disinfection as core strategies. This is also evident in the key outcomes as reported from interventions targeting the two organisms based on the reviewed studies. The identified key outcomes included antimicrobial use, resistance rates, risk reduction, adherence to contact precautions, hospital stay, and time savings. Based on the findings above, it is notable that majority (56%) of the interventions targeting *Clostridium difficile* aimed at reducing the use of antimicrobial agents. This is consistent with available evidence that demonstrates the inappropriate use of antimicrobial agents as a key risk factor for CDIs. Recent studies have shown that reducing the prescription of antimicrobials in outpatient settings can potentially reduce the incidence of CDIs in both healthcare and community settings^147, 148^. On the other hand, only 16% of the interventions targeting *Klebsiella pneumoniae* reported an impact on the use of antimicrobial agents as summarised in the findings above.

The impact of the interventions on the risk of infection was reported across the reviewed studies except for seven (39%) studies on *K. pneumoniae* that only focused on outbreak containment. Notably, the largest (97%) reduction in the absolute risk for acquiring the aforementioned organisms was reported from a multifaceted intervention targeting *Klebsiella pneumoniae* that involved surveillance, contact precautions, isolation, notification systems, and antimicrobial policies^128^. In addition, the researchers also observed that interventions targeting *Klebsiella pneumoniae* generally appear to impact more on the risk of other HCAIs when compared with interventions targeting *Clostridium difficile*. An intervention involving surveillance, contact precautions, care bundles, and environmental decontamination recorded the highest reduction in absolute risk for other HCAIs^139^. This could possibly be due to their main focus on modifying behaviours of healthcare personnel as opposed to primarily prescription behaviours as the case is evident with *Clostridium difficile* interventions.

Although most of the interventions required changes in the behaviours of healthcare personnel in breaking the transmission cycle of targeted microorganisms, there was limited evidence on the application of behaviour-based strategies to realise this objective. Only a single study incorporated Kotter’s stages of behaviour change^131^ and recorded the second largest (75%) sustained reduction in antimicrobials use over a three years period whereas the remainder of the studies were devoid of behavioural approaches.

Traditionally, health interventions such as promoting handwashing have largely relied on educational strategies to modify people’s behaviours with no attention to the underlying factors that influence people’s behaviours. On the contrary, it has become increasingly evident that providing education alone is less efficacious and does not guarantee positive behaviour change^149^. This argument is further reinforced by recent evidence that demonstrates the potential effectiveness of behavioural approaches for improved outcomes from health interventions^149–151^. As such the apparent limited application of behavioural theory in the studies reviewed under this scoping review represents a gap that warrants further exploration.

We also observed that physicians were the most involved cadre of health professionals in interventions targeting healthcare transmission of *Clostridium difficile* and *Klebsiella pneumoniae*. Almost half of the interventions in the present study involved physicians which was slightly higher than nurses (44%) whereas support staff including care workers participated in nearly one third of the interventions. In healthcare settings, physicians are amongst the least proportionate healthcare workers and their contact with patients may be less frequent compared to nurses and carers looking after patients round the clock. Consequently, it is also worth exploring whether proportionate variations in the cadres involved the above-mentioned interventions influence the key outcomes.

Finally, this review established a paucity of evidence on the application of care bundles and specialist consultations in mitigating the healthcare associated transmission of *Clostridium difficile* and *Klebsiella pneumoniae.* In addition, there was limited evidence on the effect of interventions on adherence to antimicrobial treatment protocols as well as isolation and contact precautions targeting the *Clostridium difficile* and *Klebsiella pneumoniae*.

### Limitations

There are some limitations to this scoping review. In the first place, the study population and settings of included articles were very diverse and no adjustments were undertaken to account for these differences. Secondly, the researchers did not take into account the duration of the specific interventions undertaken to mitigate *Clostridium difficile* and *Klebsiella pneumoniae* in healthcare settings. Lastly, quality assessment was not undertaken for the included studies because the main purpose of this review was basically mapping out potential sources of evidence on the topic of interest.

### Conclusions

Antimicrobial resistance represents a global threat requiring urgent measures to protect lives. Reducing the burden of AMR entails a host of multi-level approaches aimed at curbing transmission of the resistant pathogens, and optimizing the use of antibiotics. In this review, we identified the antimicrobial stewardship as well as HCAIs control interventions targeting *Clostridium difficile* and *Klebsiella pneumoniae*. These interventions include **E**ducation, **S**urveillance/**S**creening, **C**onsultations, **A**udits, **P**olicies/**P**rotocols, **E**nvironmental disinfection, **B**undles, **I**solation, and **N**otifications or alerts (ESCAPE-BIN). The key outcomes for the aforementioned interventions include antimicrobial use, cost reductions, resistance rates, risk of infection, time savings, hospital stay, as well as adherence to contact precautions and protocols. There is a further need for investigations the feasibility of behaviour-based approaches in improving adherence of health workers to interventions targeting *Clostridium difficile* and *Klebsiella pneumoniae*.

## Data Availability

The datasets generated during and/or analysed during the current study are available from the corresponding author on reasonable request through

## Appendix 1: Search strategy

### a. MEDLINE search strategy

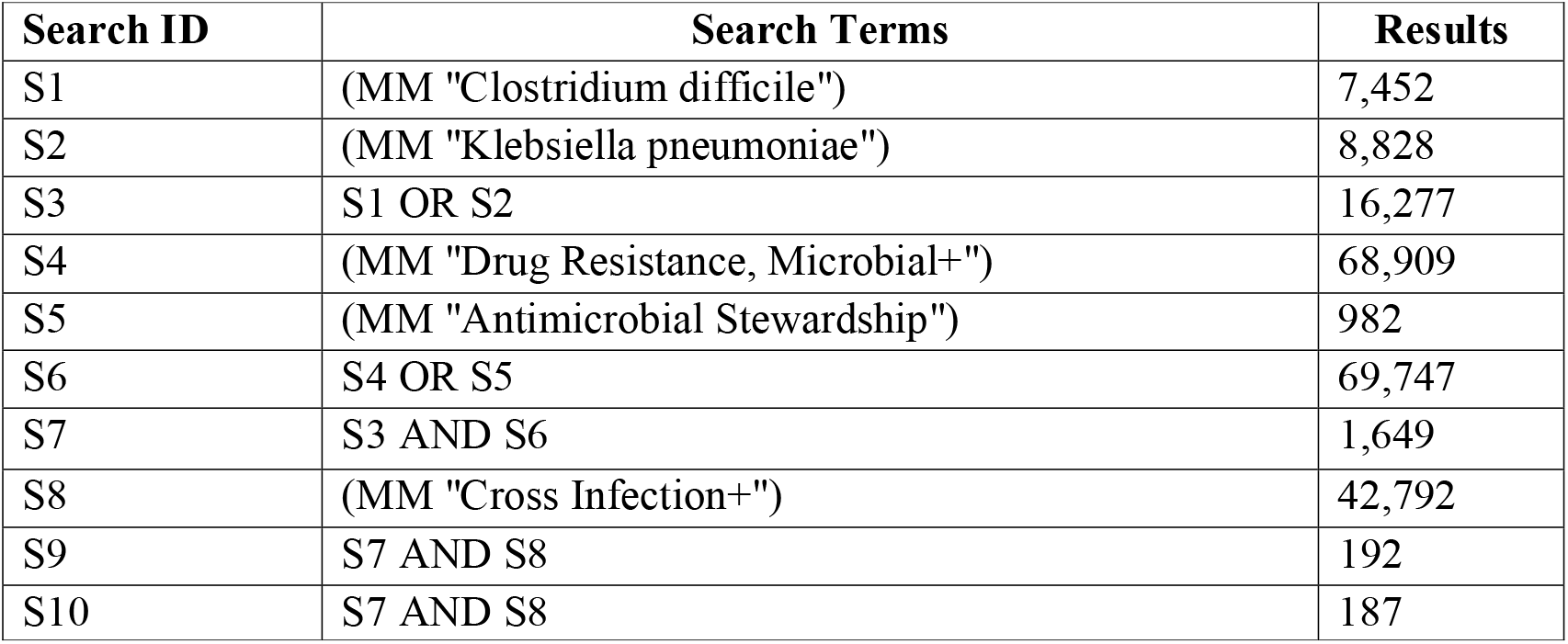

### b. CINAHL Plus

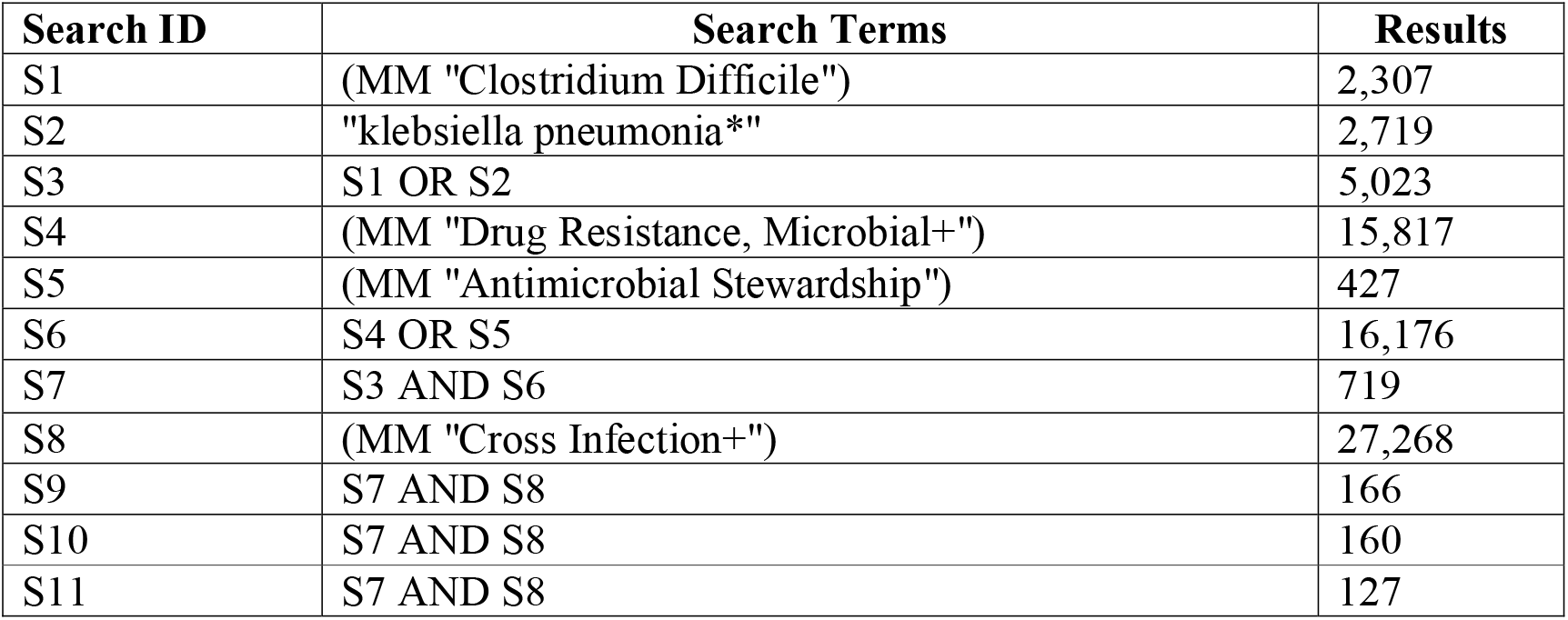

### c. Web of Science Core Collection

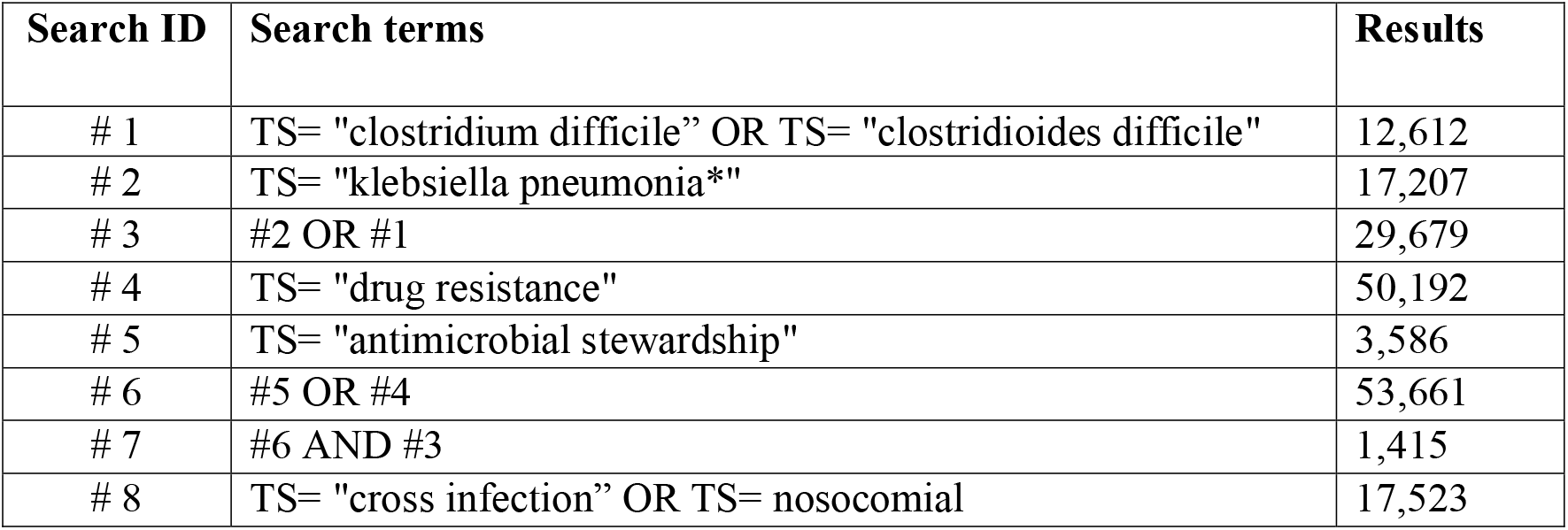
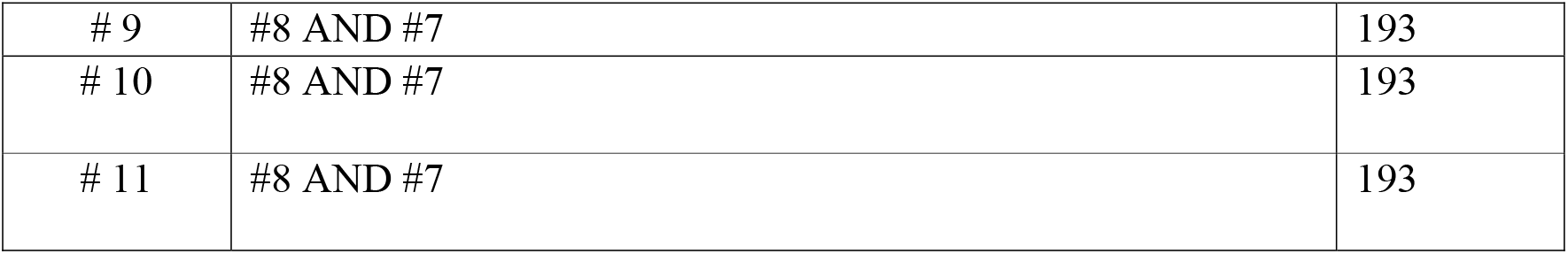

### d. PubMed

Search (“Cross Infection”[Majr]) AND ((((“Klebsiella pneumoniae”[Mesh]) OR “Clostridium difficile”[Mesh])) AND (((“Drug Resistance”[Mesh] OR “Drug Resistance, Multiple, Bacterial”[Mesh] OR “Drug Resistance, Bacterial”[Mesh] OR “Drug Resistance, Microbial”[Mesh])) OR “Antimicrobial Stewardship”[Majr])) Filters: published in the last 10 years; Humans.

## Appendix 2: Data Extraction

### a. Clostridium difficile

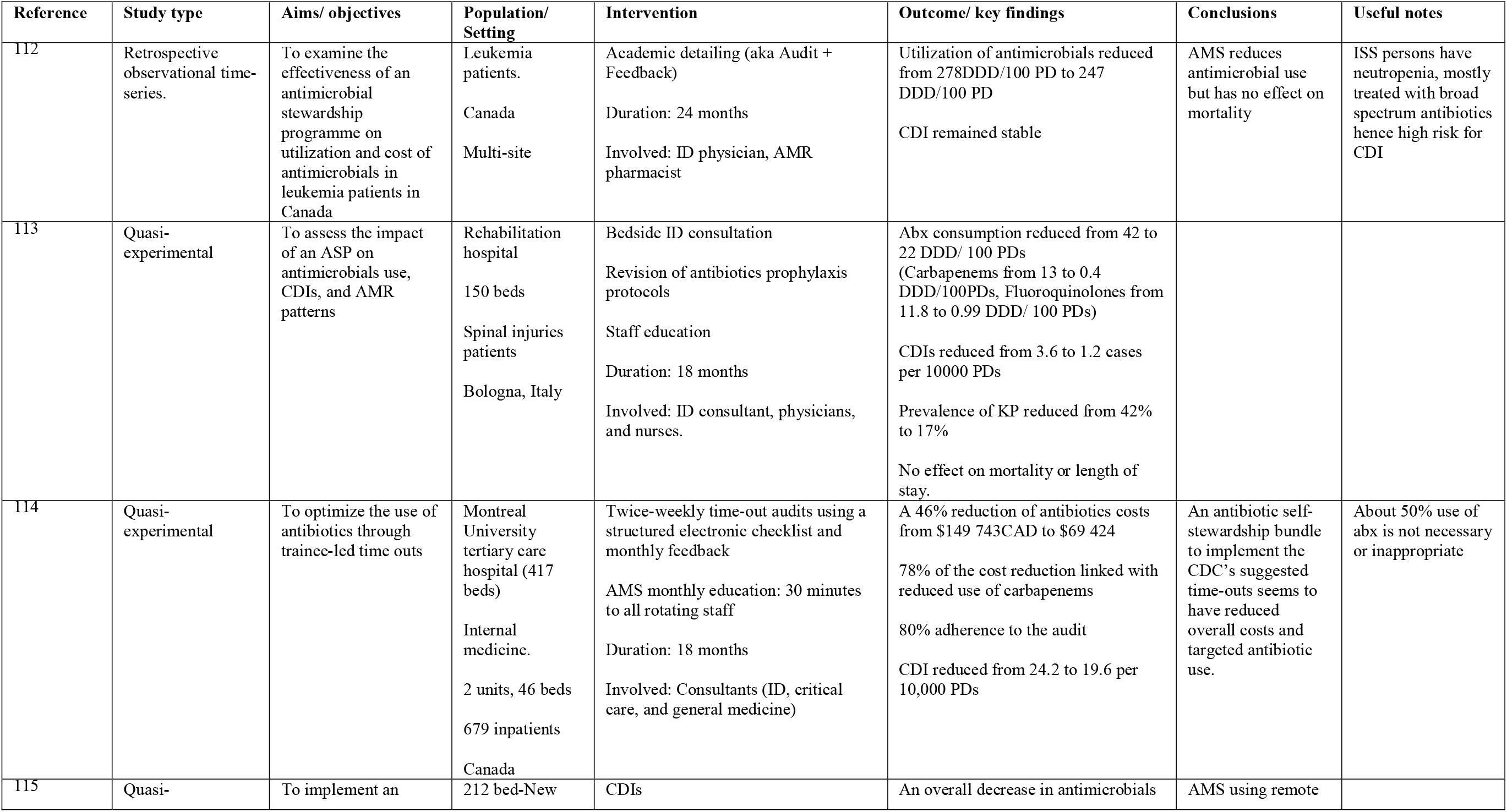
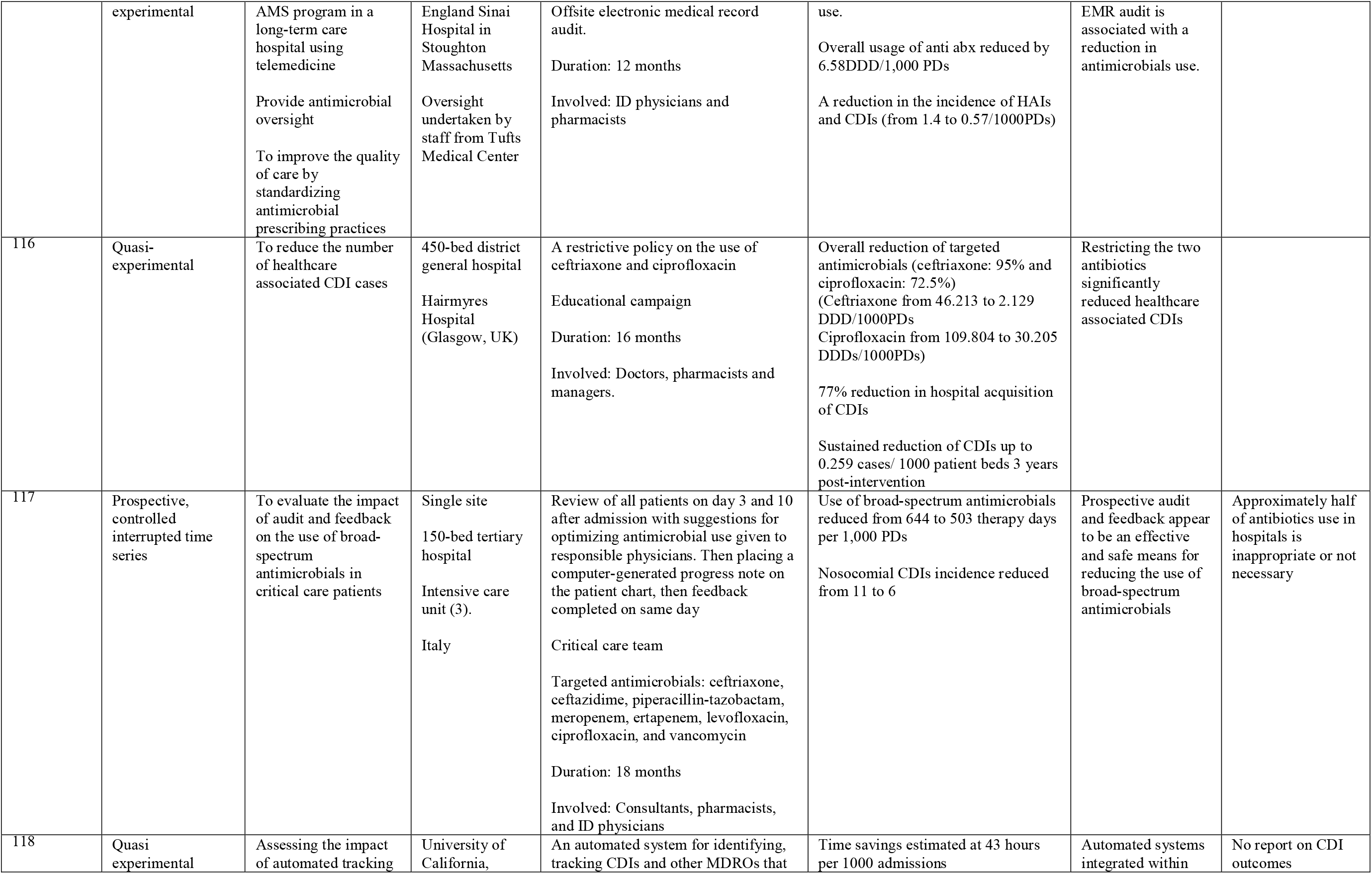
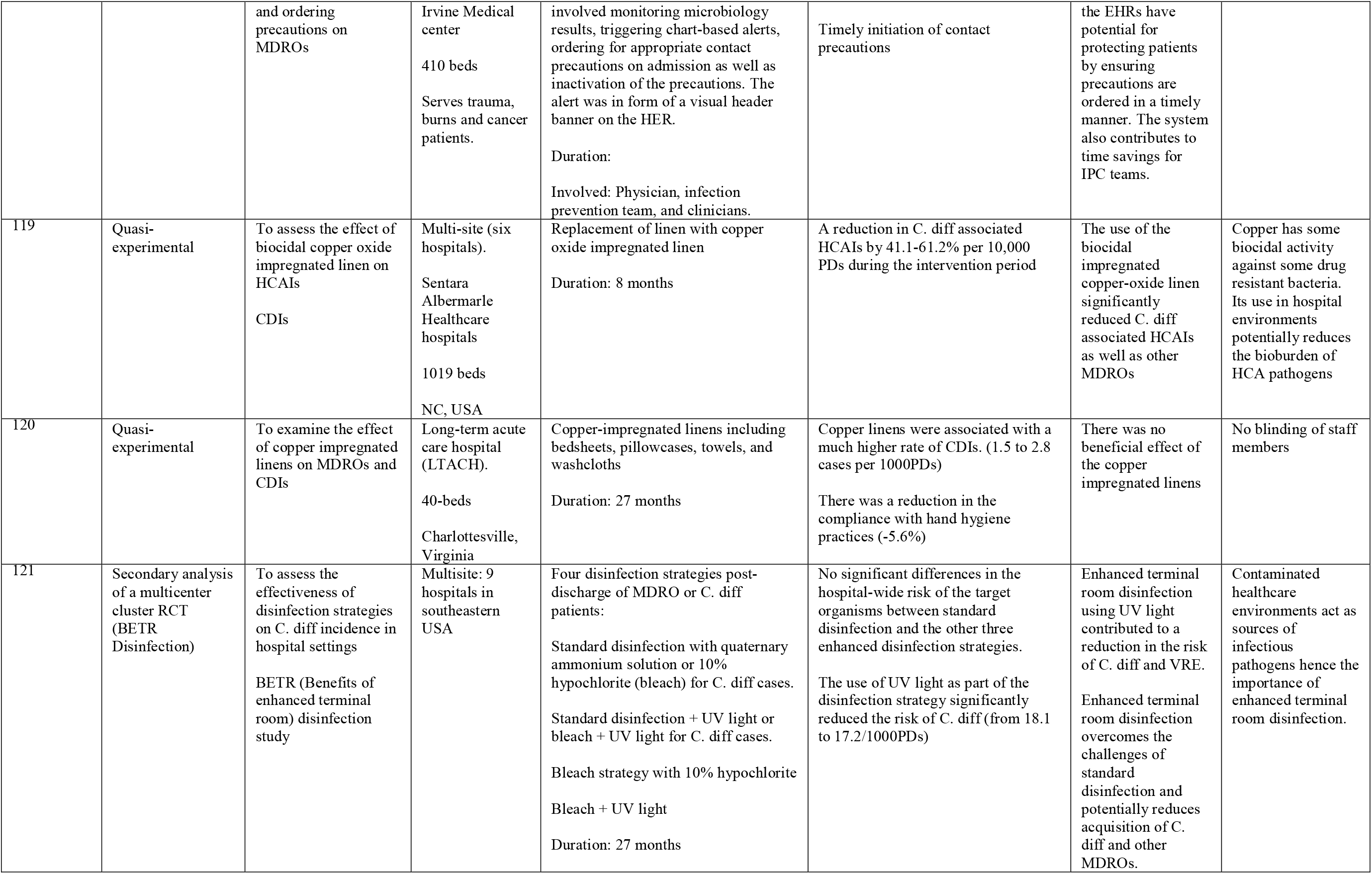
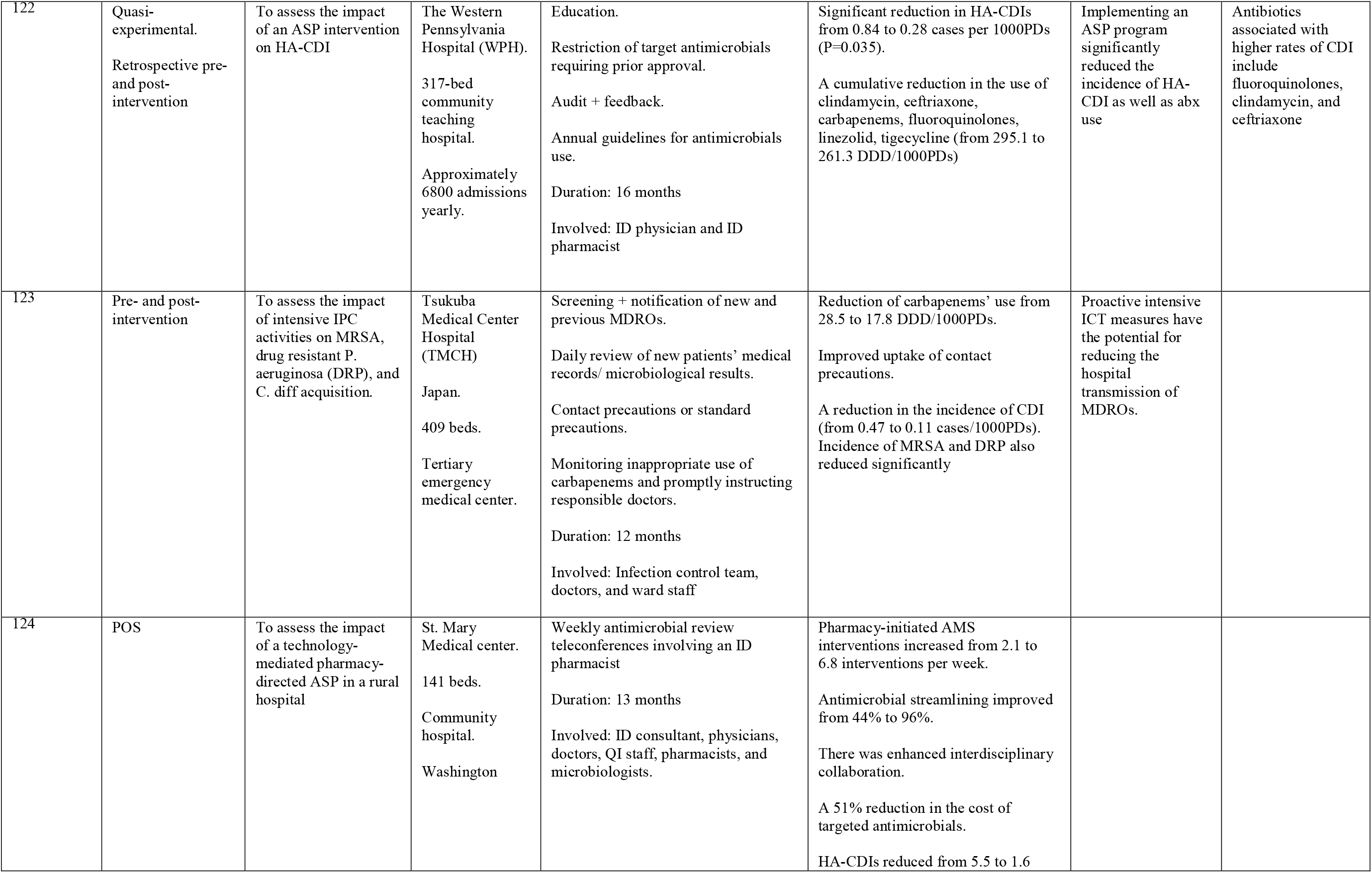
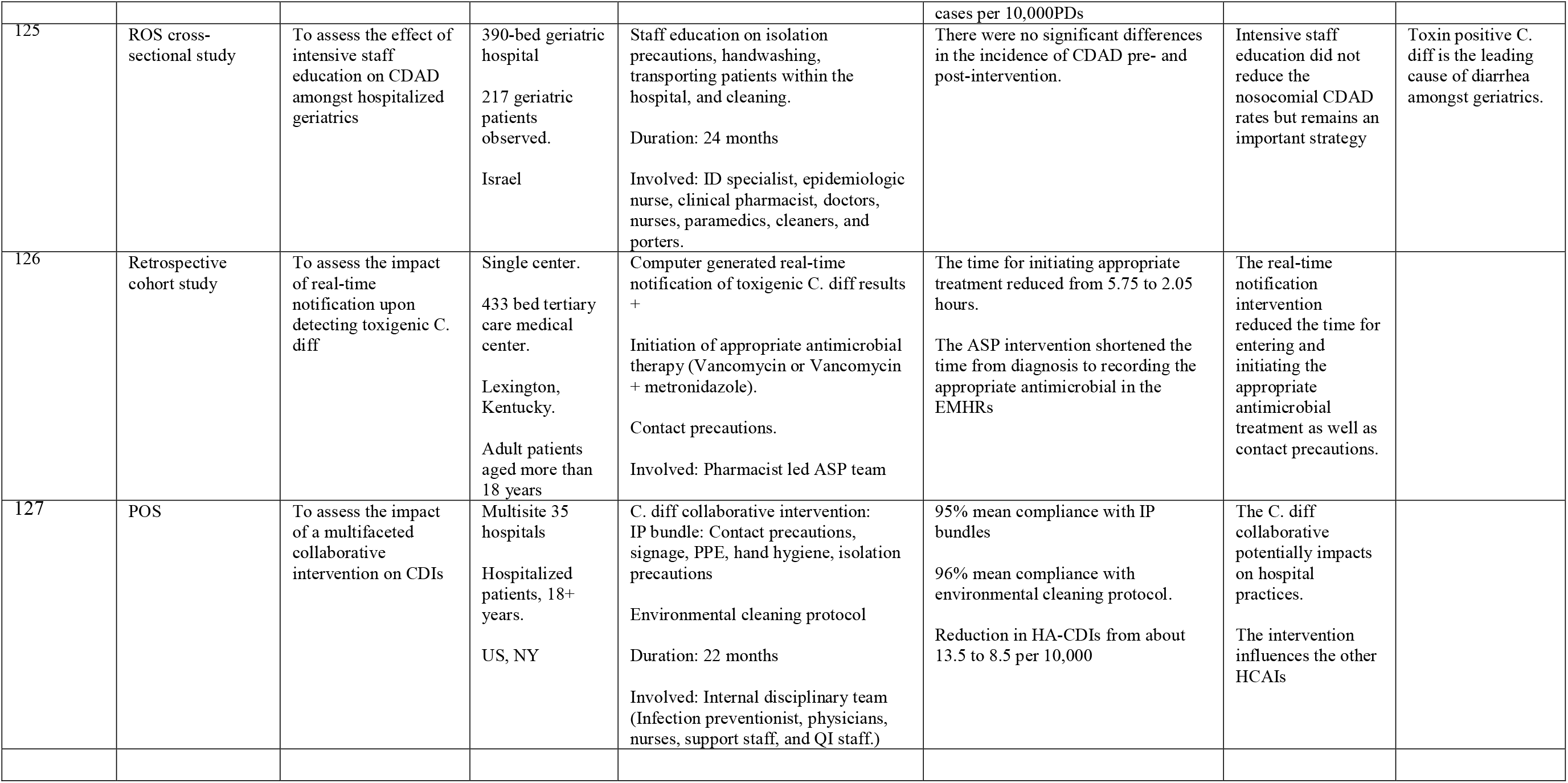

### b. Klebsiella pneumoniae

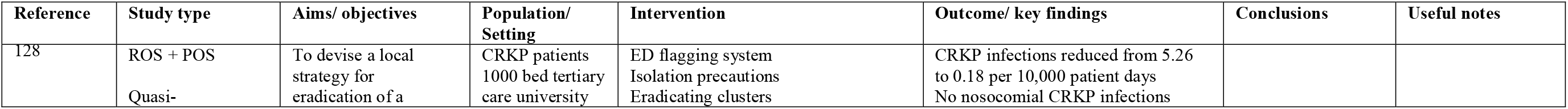
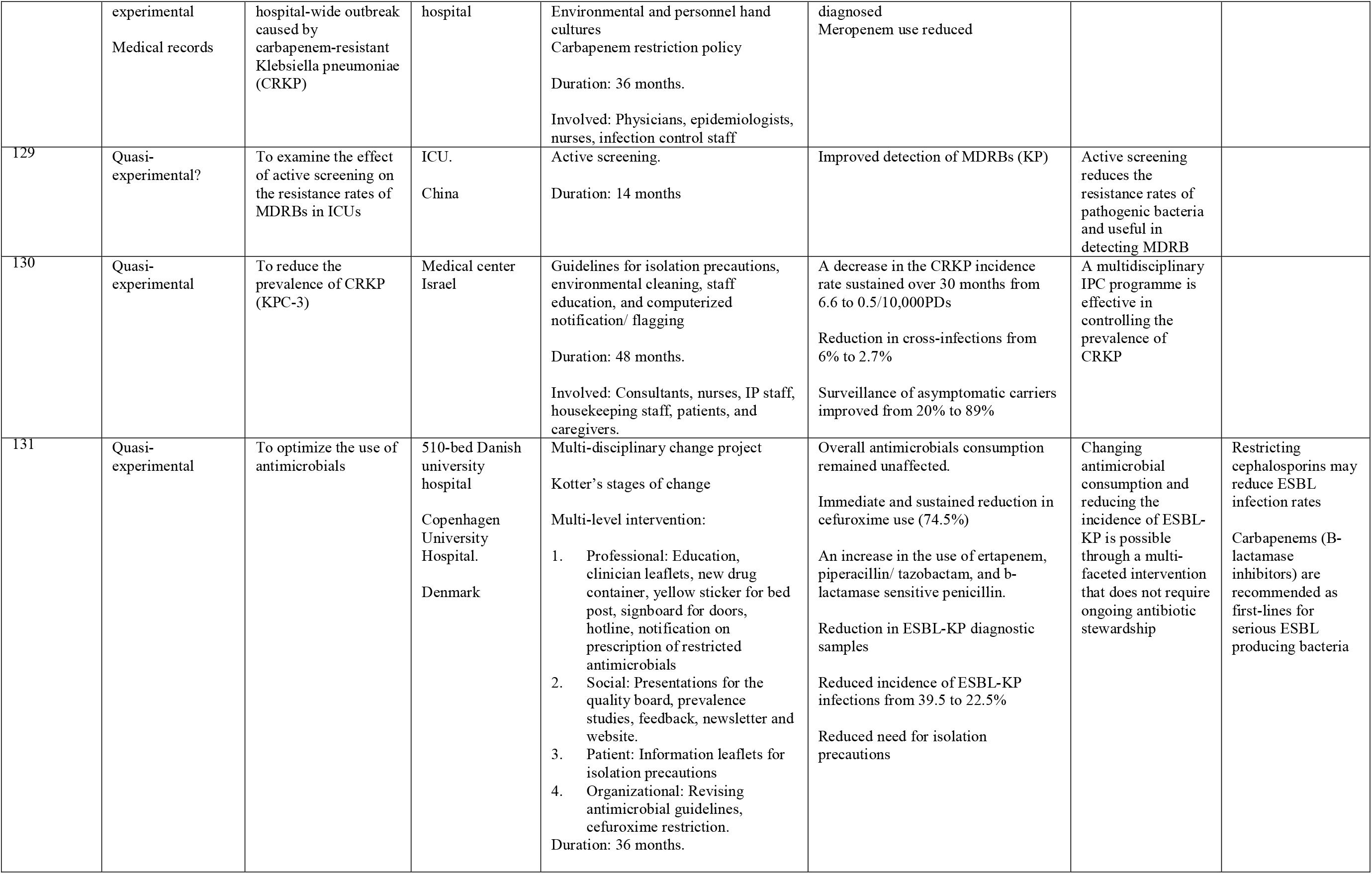
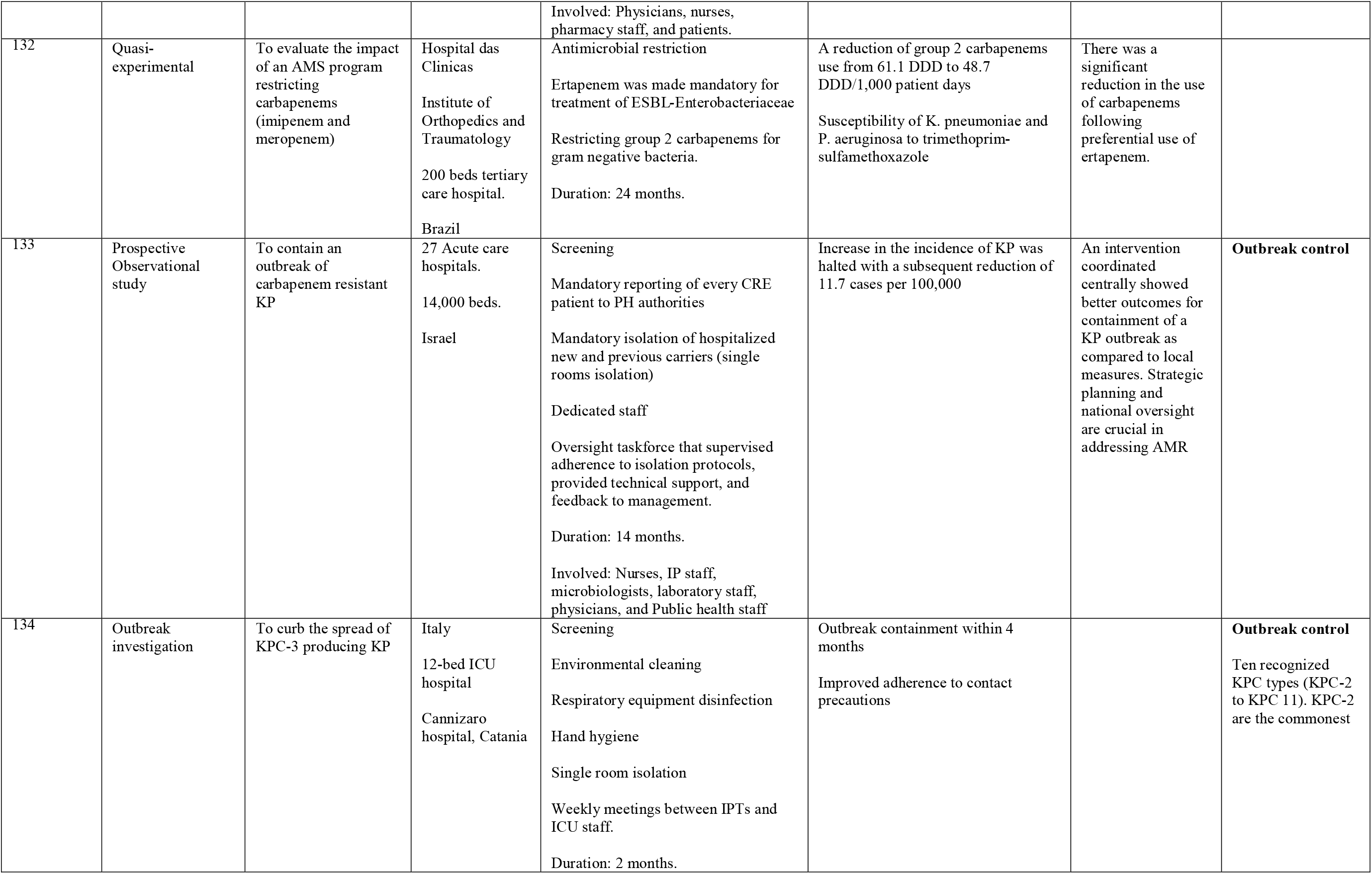
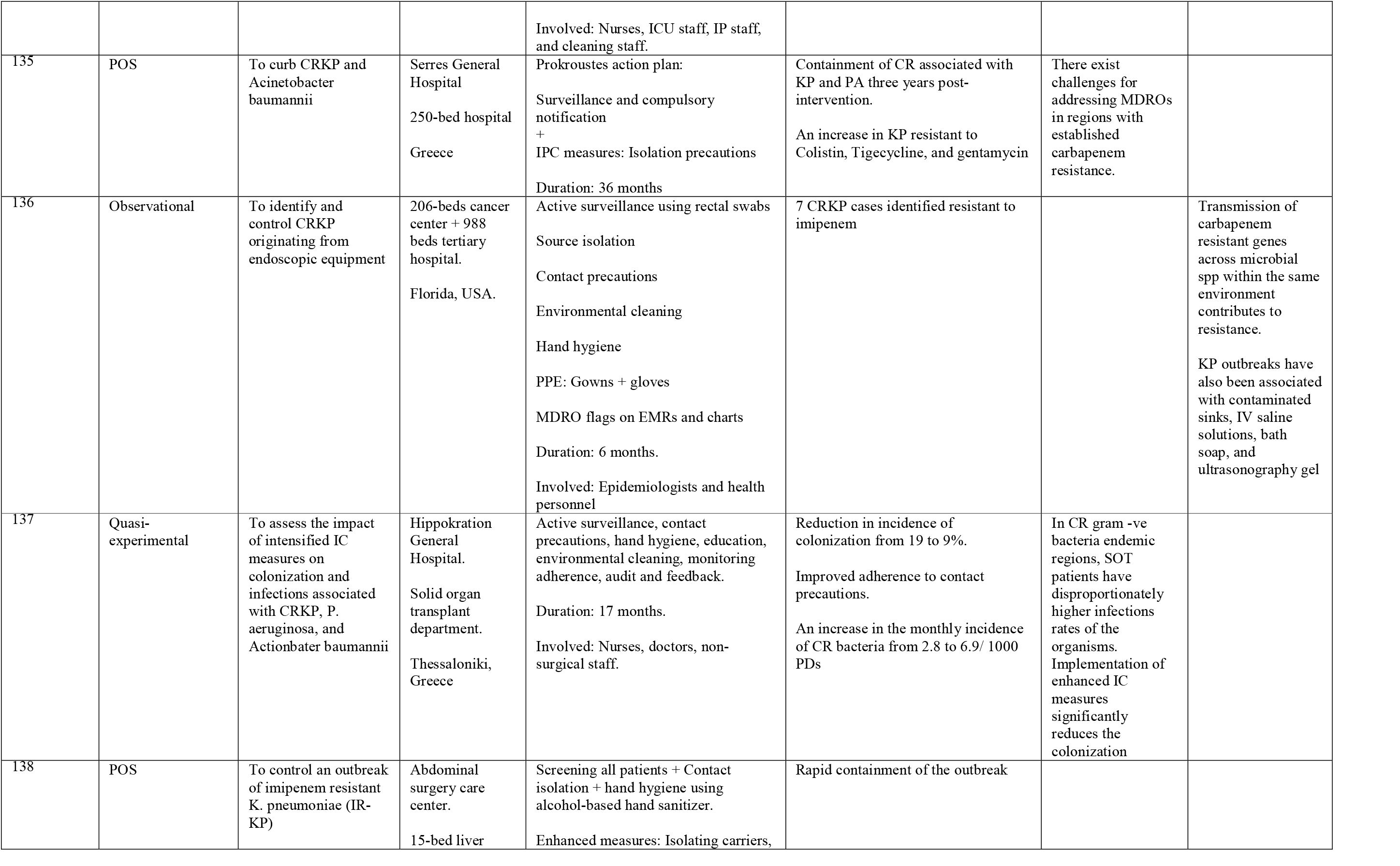
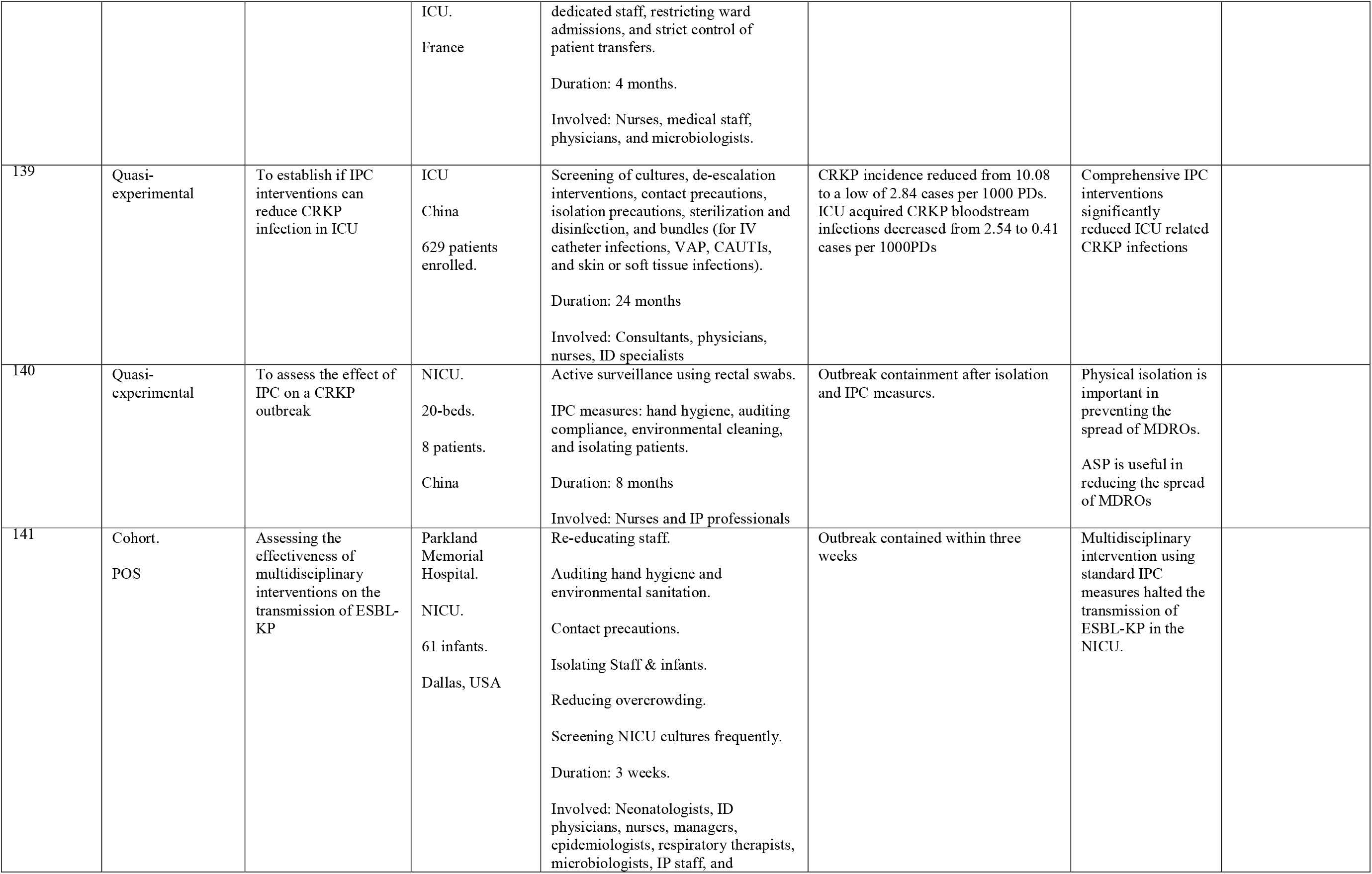
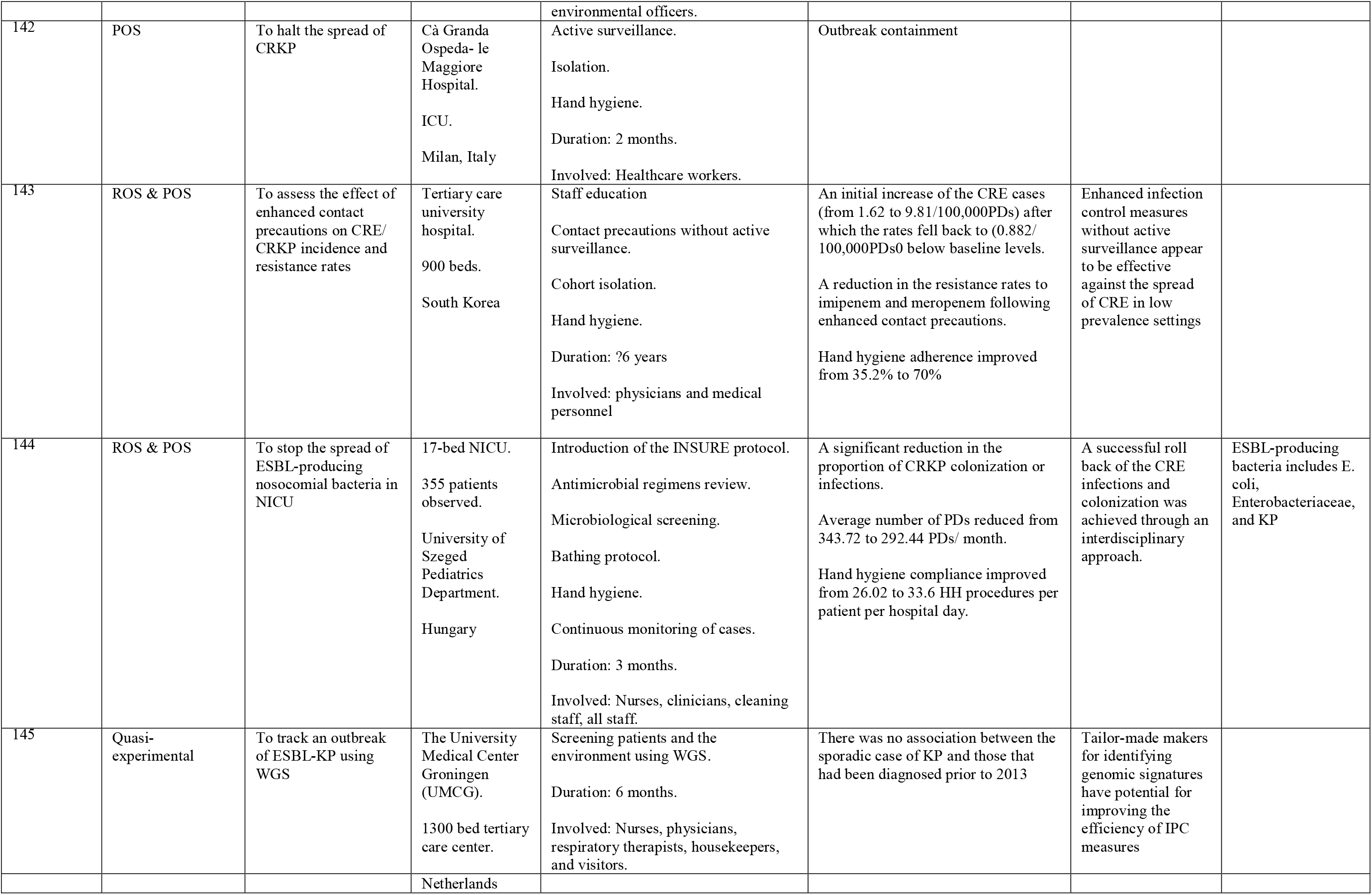

